# Effectiveness of Low-Dose Aspirin (75-150 mg) in Preventing Preeclampsia Among High-Risk Pregnant Women: A Systematic Review and Meta-Analysis of Randomized Controlled Trials

**DOI:** 10.1101/2025.03.27.25324291

**Authors:** Upma Saxena, Abhishek Lachyan, Aninda Debnath, Sunanda Gupta, Ankit Yadav, Jugal Kishore, Rishi Gupta, Sidarrth Prasad, Chanchal Goyal, Anju Sinha

**Affiliations:** Department of Obstetrics & Gynecology, VMMC & Safdarjung Hospital, New Delhi-110029; Department of Community Medicine, VMMC and Safdarjung Hospital, New Delhi; Founder and Head eStatistician Services, New Delhi; University of Texas Southwestern Medical Center, United States; Indian Council of Medical Research (ICMR), New Delhi-110029

**Keywords:** Aspirin, Bias, Meta-analysis, Preeclampsia, Prenatal care, Prevention, Randomized controlled trials

## Abstract

**Introduction:** Preeclampsia (PE) remains a significant cause of maternal and neonatal morbidity and mortality worldwide, necessitating effective prevention strategies. Low-dose aspirin has been proposed as a preventive measure for women at high risk of developing PE, given its role in modulating platelet aggregation and improving placental perfusion. However, questions regarding optimal dosage, timing, and effectiveness across diverse populations persist. This systematic review and meta-analysis aimed to synthesize evidence from randomized controlled trials (RCTs) to evaluate the efficacy of low-dose aspirin in reducing the incidence of PE.

**Methods:** A comprehensive search was conducted across databases and registers, identifying 850 records. After removing duplicates and excluding studies based on eligibility criteria, 33 RCTs were included. Data were extracted on aspirin dosage, timing of initiation, continuation duration, and PE outcomes. Meta-analysis was performed to evaluate the protective effect of aspirin, and study quality was assessed using bias scoring.

**Results:** Low-dose aspirin (75–150 mg) significantly reduced PE incidence in high-risk women. Early initiation, particularly between 12–16 weeks, demonstrated enhanced efficacy. Higher doses, such as 150 mg, were associated with better outcomes compared to lower doses. Variability among studies was noted due to differences in populations, aspirin protocols, and methodological quality. Bias assessments indicated moderate quality across most studies.

**Conclusion:** Low-dose aspirin, initiated early in pregnancy, effectively reduces PE risk in high-risk women. Findings support integrating aspirin therapy into prenatal care guidelines, with tailored dosing based on patient risk profiles. Future research should refine protocols and explore long-term maternal and neonatal outcomes.

## Introduction

Preeclampsia (PE) is a multifactorial pregnancy complication characterized by hypertension and proteinuria, affecting approximately 2-8% of pregnancies worldwide [1]. It poses significant risks to both maternal and fetal health, including maternal morbidity, preterm birth, and fetal growth restriction. In recent years, there has been a growing emphasis on identifying women at high risk for developing preeclampsia and implementing preventive strategies [2].

Low-dose aspirin (LDA), typically ranging from 75 mg to 150 mg, has emerged as a potential preventive intervention for women identified as high-risk for preeclampsia, particularly those with conditions such as chronic hypertension, diabetes, and a history of preeclampsia in previous pregnancies [3]. The mechanism by which aspirin exerts its protective effects may involve inhibition of platelet aggregation, reduction of inflammation, and improvement of placental blood flow, thereby reducing the risk of hypertensive disorders in pregnancy [4].

Several randomized controlled trials (RCTs) have evaluated the efficacy of low-dose aspirin in preventing preeclampsia in high-risk populations, yielding mixed results. While some studies have reported a significant reduction in the incidence of preeclampsia among women receiving low-dose aspirin compared to those receiving placebo, others have not demonstrated a clear benefit [5,6]. These discrepancies highlight the need for a comprehensive systematic review and meta-analysis to synthesize the available evidence and provide clearer guidance on the use of low-dose aspirin in pregnancy for the prevention of preeclampsia.

In this systematic review, we focus on assessing the efficacy and safety of low-dose aspirin for preventing preeclampsia among women identified as screened positive for high risk. The criteria for high-risk screening include a combination of clinical factors such as a history of hypertension, diabetes, kidney disease, autoimmune disorders, or other significant obstetric risk factors. Additionally, advanced screening tools like biochemical markers and uterine artery Doppler assessments were used to enhance risk identification. By consolidating data from randomized controlled trials (RCTs), this review aims to determine the impact of low-dose aspirin on reducing the incidence of preeclampsia and associated adverse outcomes, thereby informing clinical practices and supporting healthcare providers in managing high-risk pregnancies.

### Study methodology

This systematic review and meta-analysis adhered to the PRISMA (Preferred Reporting Items for Systematic Reviews and Meta-Analyses) guidelines. Only randomized controlled trials (RCTs) evaluating the use of low-dose aspirin for the prevention of preeclampsia were included. The review was registered in PROSPERO under registration number CRD42024578860.

### Inclusion and exclusion criteria

The inclusion and exclusion criteria for this systematic review were based on the PICO framework, focusing on studies where participants were high-risk pregnant women receiving low-dose aspirin (75–150 mg) for the prevention of preeclampsia. The inclusion criteria specified that the population comprised pregnant women at high or moderate risk for preeclampsia, including those with risk factors such as diabetes, chronic hypertension, autoimmune diseases, multiple gestations, chronic kidney disease, or a history of preeclampsia. The intervention considered was low-dose aspirin (75–150 mg), compared to placebo or no treatment, with the primary outcome being the development of preeclampsia. Only randomized controlled trials (RCTs) were included. The exclusion criteria ruled out studies involving pregnant women who were not at high risk for preeclampsia, studies using aspirin doses outside the 75–150 mg range, and non-randomized trials, observational studies, case reports, or reviews.

### Search Strategy

We performed a comprehensive literature search of the following electronic databases from inception to June 31, 2024: PubMed, Scopus, Web of Science and ClinicalTrials.gov. The search terms included combinations of the following keywords: “aspirin,” “low-dose aspirin,” “preeclampsia,” “pregnancy,” “randomized controlled trial,” “screened positive,” and “prevention.” The detailed search strategy for each database is provided in the supplementary material. In addition to electronic databases, we hand-searched reference lists of relevant systematic reviews, meta-analyses, and key trials for any additional studies. (Annexure 1)

|  | Inclusion Criteria | Exclusion criteria |
| --- | --- | --- |
| Population (P) | The inclusion criteria for the review encompass individuals at high and moderate risk for developing preeclampsia. High-risk factors include diabetes, chronic hypertension, autoimmune diseases, multiple gestations, chronic kidney disease, and a history of preeclampsia. Moderate-risk factors include first-time pregnancy, over 10 years between pregnancies, BMI over 30, family history of preeclampsia, age 35 or older, previous pregnancy complications, IVF conception, Black race due to systemic inequities, and lower income due to socioeconomic inequities. | Pregnant women, not at high risk for PE |
| Intervention (I) | Low dose of Aspirin (75 – 150 mg) | Any other dose of Aspirin (More than 150 mg) |
| Comparison (C) | Placebo | - |
| Outcome (O) | Development of Preeclampsia |  |
| Study Design (S) | Randomized control trial | Non randomised trials, Observational studies, Case report, Case reviews |

### Title and Abstract Screening

Two independent authors conducted the title and abstract screening of the studies retrieved from the systematic search using the predefined eligibility criteria. Any disagreements regarding the inclusion of studies for full-text review were discussed between the authors to reach a consensus. In cases where consensus could not be reached, a third author was consulted to resolve the conflict and make a final decision on whether to include the study **in** full-text screening.

### Full-Text Screening and Data Extraction

Potentially eligible full-text articles were reviewed independently by two authors to assess their suitability for inclusion. Data were then extracted from the eligible studies by the same two authors, and disagreements at any stage were resolved through discussion. If conflicts persisted, a third author made the final decision. The data extraction process was documented in a Microsoft Excel spreadsheet, where the following information was collected: author(s), location of the study, publication year, study design, sample size, number of preeclampsia cases, and intervention details. The systematic review and meta-analysis process, including the literature search, screening, and data extraction, followed the Preferred Reporting Items for Systematic Reviews and Meta-Analyses (PRISMA 2020) guidelines and was illustrated using the PRISMA flowchart.

### Risk of Bias Assessment

The quality of the included studies was assessed using the Cochrane Collaboration’s Risk of Bias 2 (RoB 2) tool. This tool evaluates bias across several domains, including the randomization process, deviations from intended interventions, missing outcome data, measurement of the outcome, and selection of the reported results. Each study was classified as having a low, some concerns, or high risk of bias. Any disagreements in quality assessment were resolved by discussion or adjudication by a third reviewer.

### Statistical analysis

The meta-analysis was conducted using a random-effects model to account for the variability between studies. The primary outcome of interest was the incidence of preeclampsia in women receiving low-dose aspirin compared to a control group. The effect sizes were reported as risk ratios (RR) with 95% confidence intervals (CI), which are appropriate for binary outcomes like preeclampsia. To ensure normality, log-transformed risk ratios were used in the forest plots. The heterogeneity between the included studies was assessed using the I^2^ statistic and Cochran’s Q test. An I^2^ value greater than 50% was considered indicative of substantial heterogeneity, signalling variability in the effect sizes across the studies due to factors beyond random error.

Subgroup analyses were performed to further investigate potential sources of heterogeneity and provide a more detailed understanding of the intervention’s effects. These analyses included examining differences in aspirin dosage and comparing the effects of 75 mg, 100 mg, and 150 mg of aspirin on the incidence of preeclampsia. The timing of aspirin initiation was another key factor, with studies divided into those that initiated aspirin before 16 weeks of gestation and those that started the intervention after 16 weeks. Additionally, we explored the impact of the continuation of aspirin by comparing studies where aspirin was stopped at 36–37 weeks with those that continued aspirin until delivery. A geographical subgroup analysis was also conducted to assess whether regional differences influenced the effectiveness of aspirin in preventing preeclampsia. Meta-regression analyses were conducted to explore sources of heterogeneity and assess how study-level characteristics influenced the overall effect size. Key covariates included sample size, aspirin dosage, and year of publication, chosen for their potential to impact observed effects. A weighted linear regression model was used, with study effect size as the dependent variable and selected covariates as predictors, and the Wald chi-square test determined the significance of each covariate’s influence.

To ensure the robustness of the findings, a sensitivity analysis was performed by excluding studies identified as having a high risk of bias. Furthermore, a leave-one-out analysis was conducted to assess the stability of the overall results by sequentially removing each study and reanalysing the data. Publication bias was assessed through a funnel plot, and any potential asymmetry was tested using Egger’s test. To further address and correct for any asymmetry observed in the funnel plot, the trim-and-fill method was applied, estimating and imputing potentially missing studies to provide an adjusted and more balanced overall effect size by accounting for unpublished or missing studies. We used STATA 18 for analysis purposes.

### Software used

#### Methods (Literature Screening)

We used *Rayyan*, a web-based software tool, to streamline the screening of titles and abstracts for eligibility.

#### Methods (Statistical Analysis)

Forest plots and other graphical representations of data were generated using *STATA 18*.

### Ethical Statement

An ethical review was not required for this study as it exclusively used data available from previously published sources.

## Results

A total of 780 relevant articles were retrieved from various databases and registers, with an additional 70 articles identified from the Clinical Trial Register, resulting in 850 records. After removing 495 duplicate records, 285 articles remained for screening. During the screening process, 220 articles were excluded based on titles and abstracts. The full text of 65 articles was sought, but 5 reports were not retrieved. After assessing 60 full-text articles for eligibility, 33 studies were included in the final meta-analysis. Key reasons for exclusion included incorrect aspirin dosage (n = 8), wrong population (n = 10), absence of the outcome variable (n = 7), and foreign language (n = 2). Ultimately, 33 studies were included in the review. (Figure 1)

**Figure 1:**
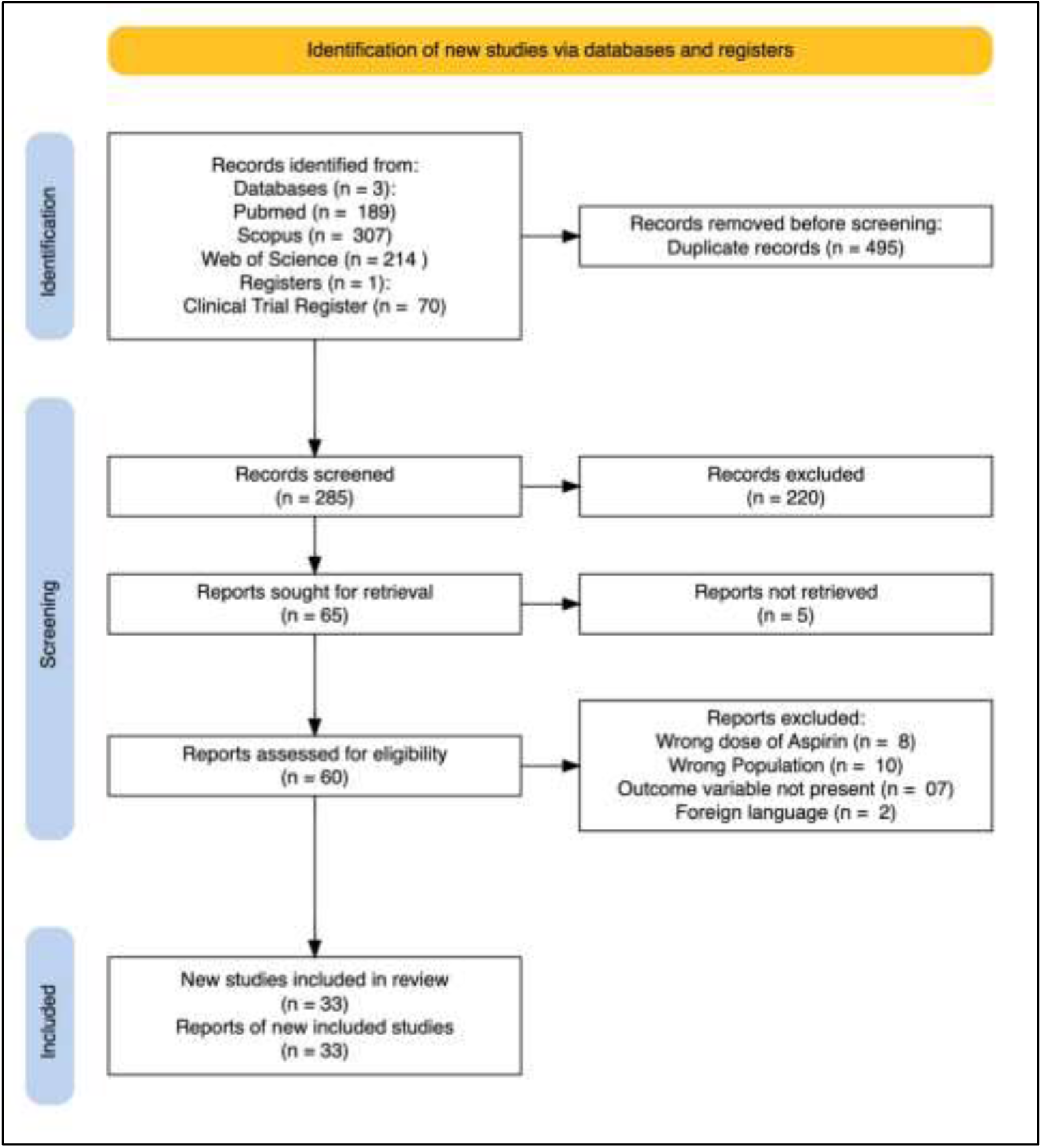
PRISMA flow diagram of inclusion of studies

**Table 1:** Details of the studies included in the analysis.

| **Sl no** | **Author (Last name et al.)** | **Continent** | **Dose of aspirin** | **aspirin was started** | **Aspirin was continued** | **Participants analysed in Aspirin group** | **Aspirin with PE** | **Participants analysed in Placebo group** | **Placebo with PE** |
| --- | --- | --- | --- | --- | --- | --- | --- | --- | --- |
| **1** | Abdi et al. (2020) [7] | Asia | 80 | 12-15 weeks | 36 week/ Until delivery | 43 | 27 | 43 | 38 |
| **2** | Ali et al (2018) [8] | Asia | 100 | 16 weeks | 36 weeks | 25 | 4 | 25 | 10 |
| **3** | Ayala et al (2012) [9] | Europes | 100 | 12-16 weeks | Until delivery | 176 | 11 | 174 | 22 |
| **4** | Bakhti et al. (2011) [10] | Africa | 100 | 8-10 weeks | 36 weeks of gestation | 82 | 0 | 82 | 4 |
| **5** | Byaruhanga et al.(1998) [11] | Africa | 75 | 20-28 weeks | 36 weeks of gestation | 113 | 17 | 117 | 23 |
| **6** | Chiaffarino et al.(2004) [12] | Europe | 100 | <14 weeks | Until delivery | 16 | 5 | 19 | 7 |
| **7** | Davies et al (1995) [13] | Europe | 75 | 18 weeks | Until delivery | 58 | 5 | 60 | 7 |
| **8** | Ebrashy et al.(2005) [14] | Africa | 75 | 14-16 weeks | Until delivery | 74 | 26 | 65 | 40 |
| **9** | Grab et al. (2000) [15] | Europe | 100 | 20 weeks | Until Delivery | 22 | 3 | 21 | 2 |
| **10** | Gu et al. (2020) [16] | Asia | 75 | 12 weeks | Until delivery | 271 | 26 | 284 | 51 |
| **11** | Haapsamo et al. (2010) [17] | Europe | 100 | On the first day of gonadotrophin stimulation | Until delivery | 52 | 4 | 55 | 4 |
| **12** | Harrington et al.(2000) [18] | Europe | 100 | 20 weeks | 37 weeks of gestation | 58 | 6 | 103 | 9 |
| **13** | Hermida (2002) [19] | Europe | 100 | 12-16 weeks | Until delivery | 174 | 27 | 167 | 24 |
| **14** | Hermida et al (1997) [20] | Europe | 100 | 12-16 weeks | Until delivery | 50 | 3 | 50 | 7 |
| **15** | Huai J et al. (2021) [21] | Asia | 100 | 12-20 weeks | 34 weeks of gestation | 139 | 6 | 144 | 16 |
| **16** | Jamal et al. (2012) [22] | Asia | 80 | 12 weeks | Until delivery | 35 | 2 | 35 | 4 |
| **17** | Khazardoost et al (2014) [23] | Asia | 80 | 15-18 weeks | 32 weeks of gestation | 65 | 5 | 68 | 8 |
| **18** | Lin Li et al (2022) [24] | Asia | 100 | 12-20 weeks | 34 weeks of gestation | 464 | 78 | 434 | 74 |
| **19** | Liu et al.(2016) [25] | Asia | 100 | 4-15 weeks | Until delivery | 50 | 3 | 48 | 10 |
| **20** | McCowan et al (1999) [26] | Oceania | 100 | 24-36 weeks | Taken treatment for more than 14 days | 32 | 7 | 33 | 4 |
| **21** | McParland et al. (1990) [27] | Europe | 75 | 24 weeks of gestation | Until delivery | 48 | 6 | 52 | 13 |
| **22** | Mone et al. (2018) [28] | Europe | 75 | 11-14 weks | 36 weeks of gestation | 192 | 8 | 354 | 7 |
| **23** | Morris et al (1996) [29] | Oceania | 100 | 18 weeks gestation | Until delivery | 52 | 4 | 50 | 7 |
| **24** | Odibo et al. (2015) [30] | North America | 81 | 11-13 weeks | Until delivery | 16 | 3 | 14 | 3 |
| **25** | Poon et al (2017) [31] | Europe | 150 | 11-14 weks | 36 weeks of gestation | 798 | 13 | 822 | 35 |
| **26** | Rogers et al. (1999) [32] | Asia | 80 | 22 weeks | Until Delivery | 132 | 3 | 83 | 7 |
| **27** | Rolnik et al.(2017) [33] | Europe | 150 | 11-14 weks | 36 weeks of gestation | 785 | 87 | 806 | 94 |
| **28** | Scazzocchio et al (2017) [34] | Europe | 150 | 11-14 weks | 28 weeks of gestation | 80 | 5 | 75 | 4 |
| **29** | Schiff et al (1989) [35] | Asia | 100 | 28-29 weeks | 38+4 weeks of gestation | 34 | 1 | 31 | 7 |
| **30** | Schrocksnadel et al (1992) [36] | Europe | 80 | 28-32 Weeks | 37 weeks of gestation | 22 | 0 | 19 | 6 |
| **31** | Talari et al (2014) [37] | Asia | 80 | 12-16 Weeks | - | 40 | 1 | 40 | 9 |
| **32** | Villa et al. (2013) [38] | Europe | 100 | 12-13 | Until Delivery | 61 | 8 | 60 | 11 |
| **33** | Yu et al. (2003) [39] | Multicontinental | 150 | 23 weeks | 36 weeks of gestation | 276 | 49 | 278 | 52 |

**Figure 2:**
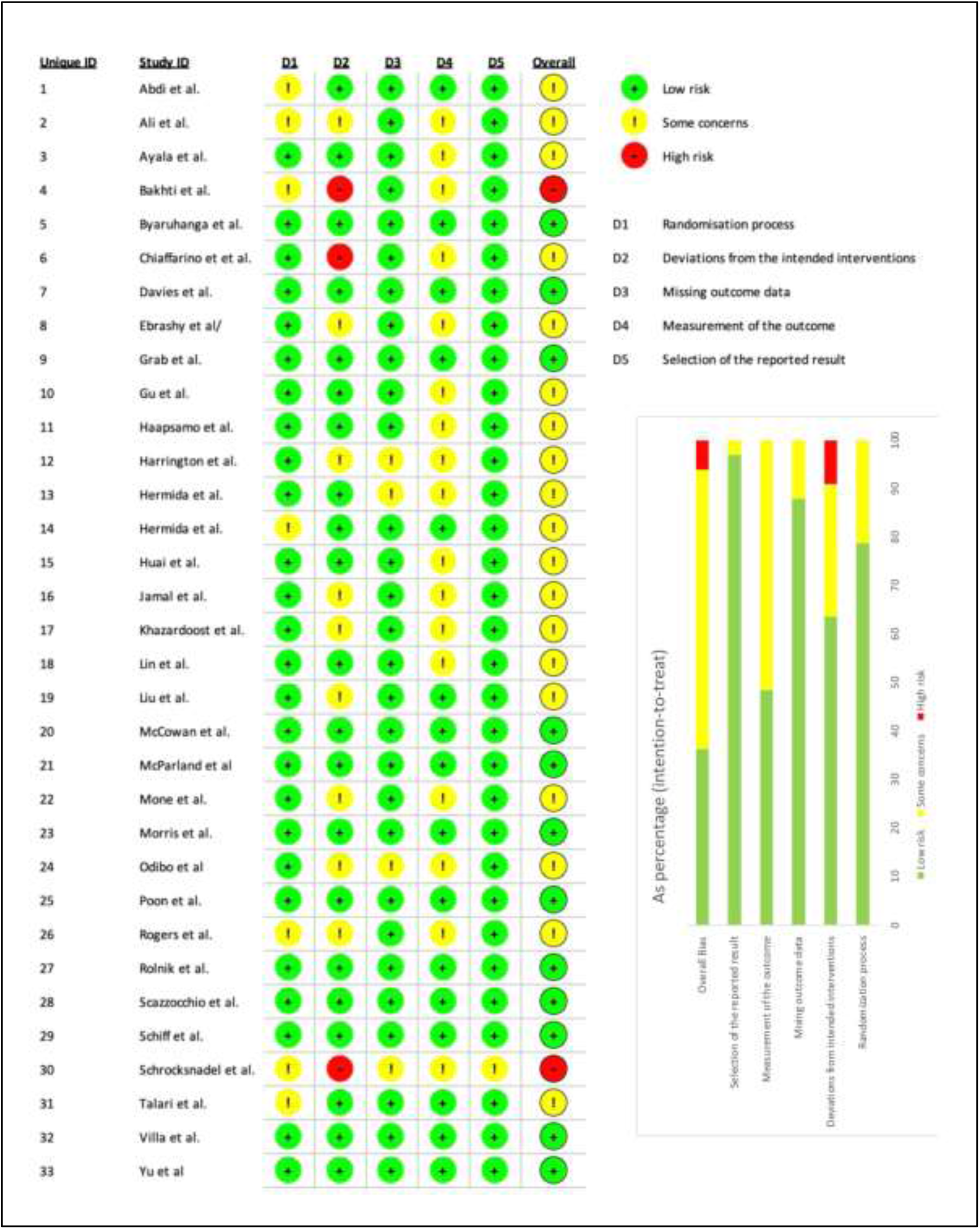
Bias scoring of the studies included in final analysis

## Discussion

The findings of this systematic review and meta-analysis offer compelling evidence for the use of low-dose aspirin (75 mg - 150 mg) as a preventive measure against preeclampsia in women screened as high-risk. The data synthesized from various randomized controlled trials (RCTs) underscores the importance of early intervention and appropriate dosing in mitigating the risk of this potentially severe pregnancy complication.

### 1. Efficacy of Low-Dose Aspirin

The analysis reveals a significant reduction in preeclampsia rates among women receiving low-dose aspirin compared to those on placebo. The studies included in this review commenced aspirin treatment at various gestational ages, predominantly between 12 and 16 weeks. The results support the notion that early initiation of aspirin is crucial for maximizing its protective effects. Notably, trials such as Ayala et al. (2012) [9] and Lin Li et al. (2022) [24] reported marked decreases in preeclampsia incidence, reinforcing the importance of starting therapy in the first trimester.

Dosing also appears to play a critical role in the efficacy of aspirin. Our review indicates that higher doses, particularly 100 mg and 150 mg, are associated with lower rates of preeclampsia compared to 75 mg. For instance, Poon et al. (2017) [31] demonstrated that 150 mg of aspirin significantly reduced preeclampsia incidence, suggesting a potential dose-response relationship. This finding aligns with previous research indicating that larger doses might provide enhanced protection for women with multiple risk factors.

### 2. Variability Among Studies

Despite the overall positive outcomes, there is notable variability among the studies included in this review. The included trials varied in their participant demographics, definitions of preeclampsia, and methodological quality. For example, some studies involved small sample sizes or lacked detailed descriptions of blinding and randomization processes, raising concerns about bias and the generalizability of findings.

Additionally, geographical differences in the studies may have influenced outcomes. Trials conducted in Asia and Europe provided diverse insights, but variations in population characteristics, healthcare systems, and environmental factors could contribute to different preeclampsia rates. Such heterogeneity highlights the need for more consistent protocols in future studies to ensure that findings are robust and applicable across diverse populations.

### 3. Mechanisms of Action

The protective effect of low-dose aspirin against preeclampsia is believed to stem from its ability to inhibit platelet aggregation and modulate inflammatory processes. By reducing thromboxane A2 levels and increasing prostacyclin availability, aspirin can improve placental perfusion and reduce the risk of endothelial dysfunction, which is central to the pathophysiology of preeclampsia. Understanding these mechanisms provides a biological rationale for the observed benefits of aspirin therapy in high-risk populations.

### 4. Clinical Implications and Recommendations

The implications of these findings are significant for clinical practice. The use of low-dose aspirin in women identified as high-risk for preeclampsia should be integrated into routine prenatal care, particularly for those with prior adverse pregnancy outcomes, chronic hypertension, or multiple gestational factors. Educating patients on the benefits and potential risks associated with aspirin therapy is vital, ensuring informed decision-making regarding its use.

Healthcare providers should adopt a tailored approach, considering individual patient risk profiles when recommending aspirin. For example, women with a history of severe preeclampsia may benefit from higher doses, while those with fewer risk factors might find lower doses adequate. Continuous monitoring of patient outcomes and adherence to guidelines will be essential for maximizing the benefits of aspirin therapy.

### 5. Future Research Directions

While this review provides a robust synthesis of current evidence, further research is warranted to refine guidelines regarding aspirin use in pregnancy. Future studies should aim to:

- Investigate the optimal timing and duration of aspirin therapy, particularly in diverse populations.
- Explore the long-term effects of low-dose aspirin on maternal and neonatal outcomes beyond preeclampsia prevention, including potential impacts on long-term cardiovascular health.
- Assess the safety and efficacy of low-dose aspirin in women with preexisting conditions or comorbidities, ensuring comprehensive risk assessments.

## 6. Limitations of the Review

This systematic review has limitations that must be acknowledged. The heterogeneity of study designs, including variations in aspirin dosage, initiation timing, and participant characteristics, may impact the reliability of pooled outcomes. Moreover, the lack of standardized definitions for preeclampsia across studies complicates the comparison of results. Bias related to publication status and selective reporting may also skew the overall findings.

Future meta-analyses should prioritize high-quality studies with standardized protocols to enhance the reliability of conclusions drawn regarding the use of low-dose aspirin in pregnancy.

## 7. Conclusion

In summary, low-dose aspirin demonstrates a significant protective effect against preeclampsia in women screened as high-risk, particularly when initiated early in pregnancy and at higher doses. These findings underscore the importance of proactive management strategies in obstetric care. As our understanding of preeclampsia evolves, integrating low-dose aspirin into preventive protocols could play a pivotal role in improving maternal and neonatal health outcomes. Ongoing research will be crucial in refining treatment guidelines and enhancing our ability to prevent this serious condition.

**Figure 3:**
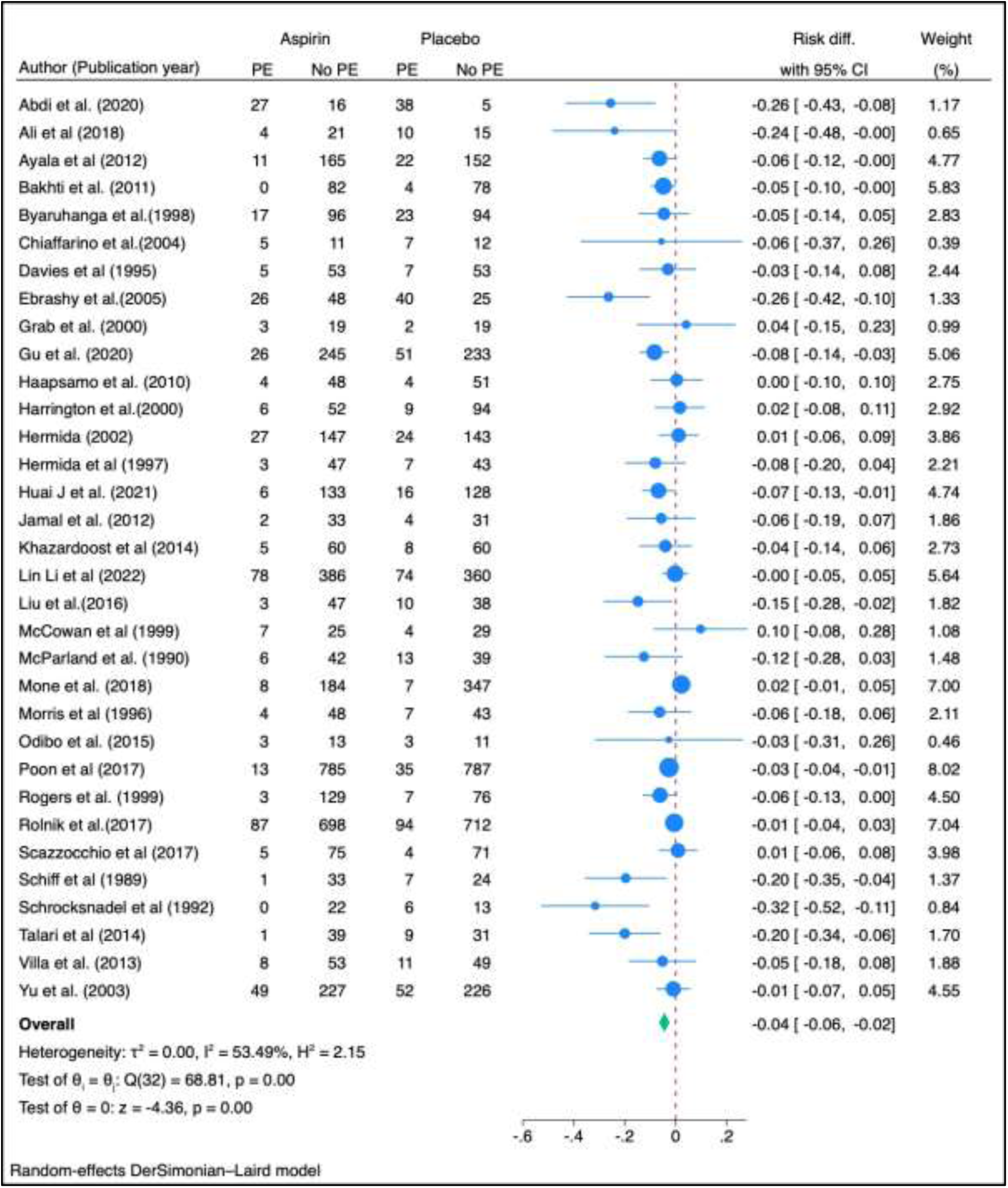
A total of 33 studies were included in the meta-analysis, examining the effect of low-dose aspirin on the prevention of preeclampsia (PE) in high-risk pregnant women. The pooled analysis, using a random-effects model, demonstrated a statistically significant reduction in the risk of preeclampsia associated with aspirin use. The overall risk difference (RD) was −0.04 (95% CI: −0.06 to −0.02, p = 0.001), indicating a 4% absolute reduction in the risk of developing preeclampsia in the aspirin group compared to the placebo group. The heterogeneity among the studies was moderate, with an I^2^ of 53.49%, suggesting some variability between study outcomes. However, the between-study variance was low in absolute terms (τ^2^ = 0.00), and the Q-test (p = 0.001) indicated significant heterogeneity across studies. These findings suggest that low-dose aspirin is effective in reducing the risk of preeclampsia in high-risk pregnancies, with consistent results across a diverse range of studies, despite some degree of heterogeneity.

**Figure 4:**
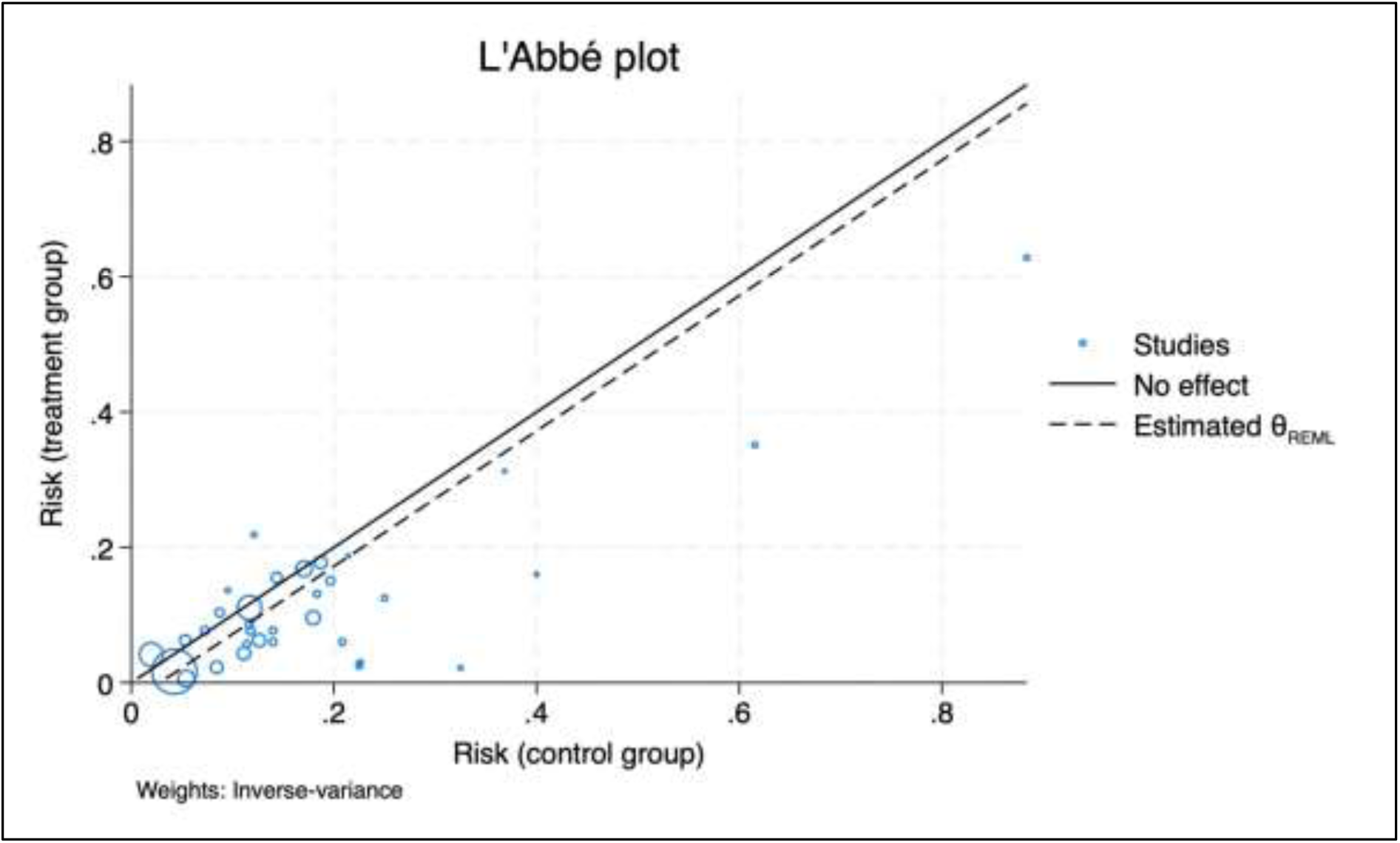
The L’Abbé plot visually supports the conclusion that low-dose aspirin is associated with a reduction in the risk of preeclampsia in high-risk pregnancies. The clustering of studies below the no effect line demonstrates a consistent beneficial effect of aspirin across most of the included studies.

**Figure 5:**
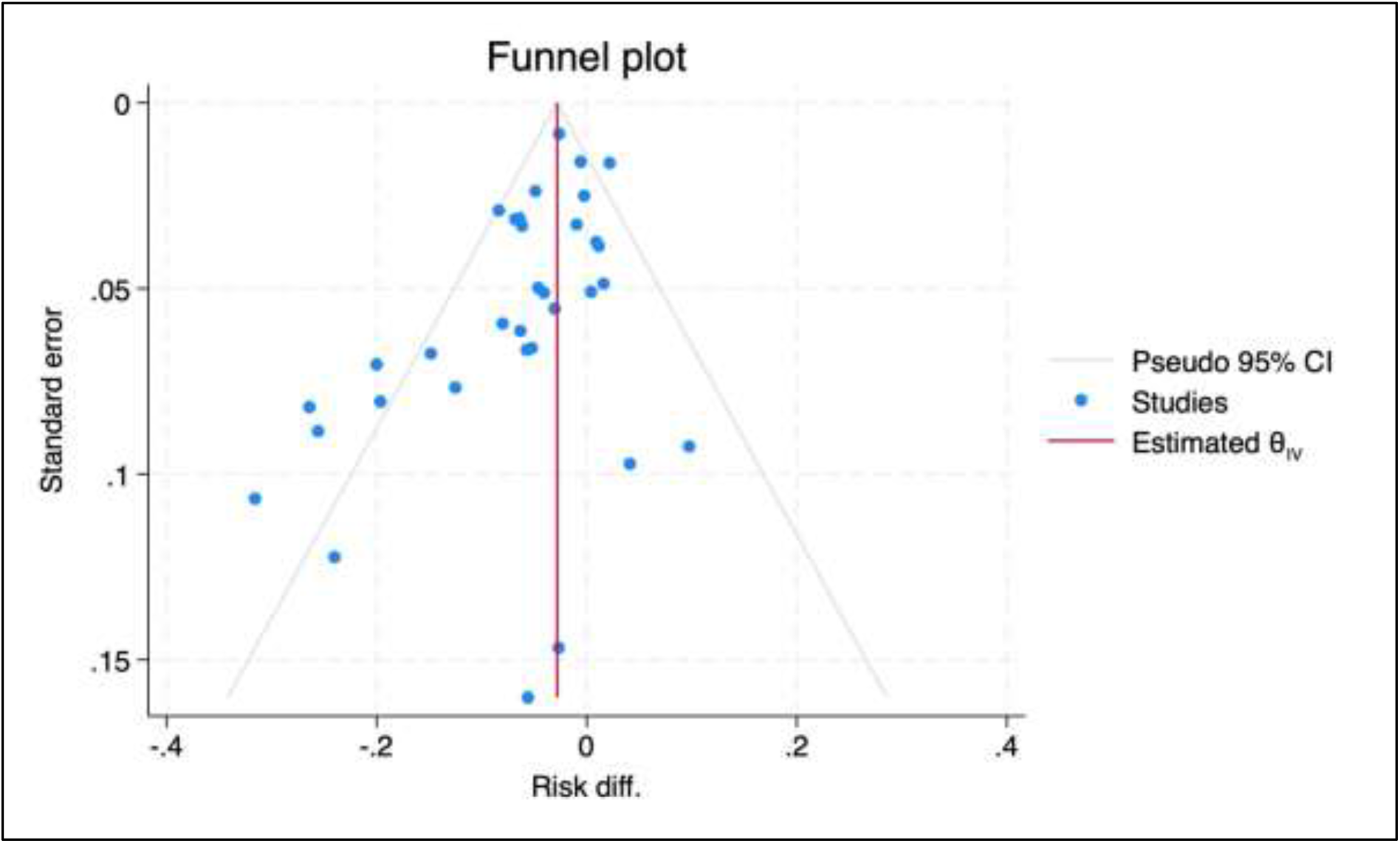
Egger’s test for small-study effects provided statistical support for the presence of publication bias, with a significant p-value of 0.05. The negative beta coefficient (−1.31) indicated that smaller studies were more likely to report larger negative effects, further suggesting a bias towards publishing studies that showed more substantial benefits of the intervention. The funnel plot for risk differences initially showed some asymmetry, with a noticeable skew to the left. The trim-and-fill method identified five potentially missing studies on the right side of the funnel plot. These imputed studies were added to correct for the suspected bias. After imputing these studies, the adjusted overall risk difference shifted, still indicating a protective effect of aspirin but with a reduced magnitude. This suggests that while publication bias likely inflated the observed effect size, the overall conclusion of aspirin’s protective effect remains consistent, though slightly diminished.

**Figure 6:**
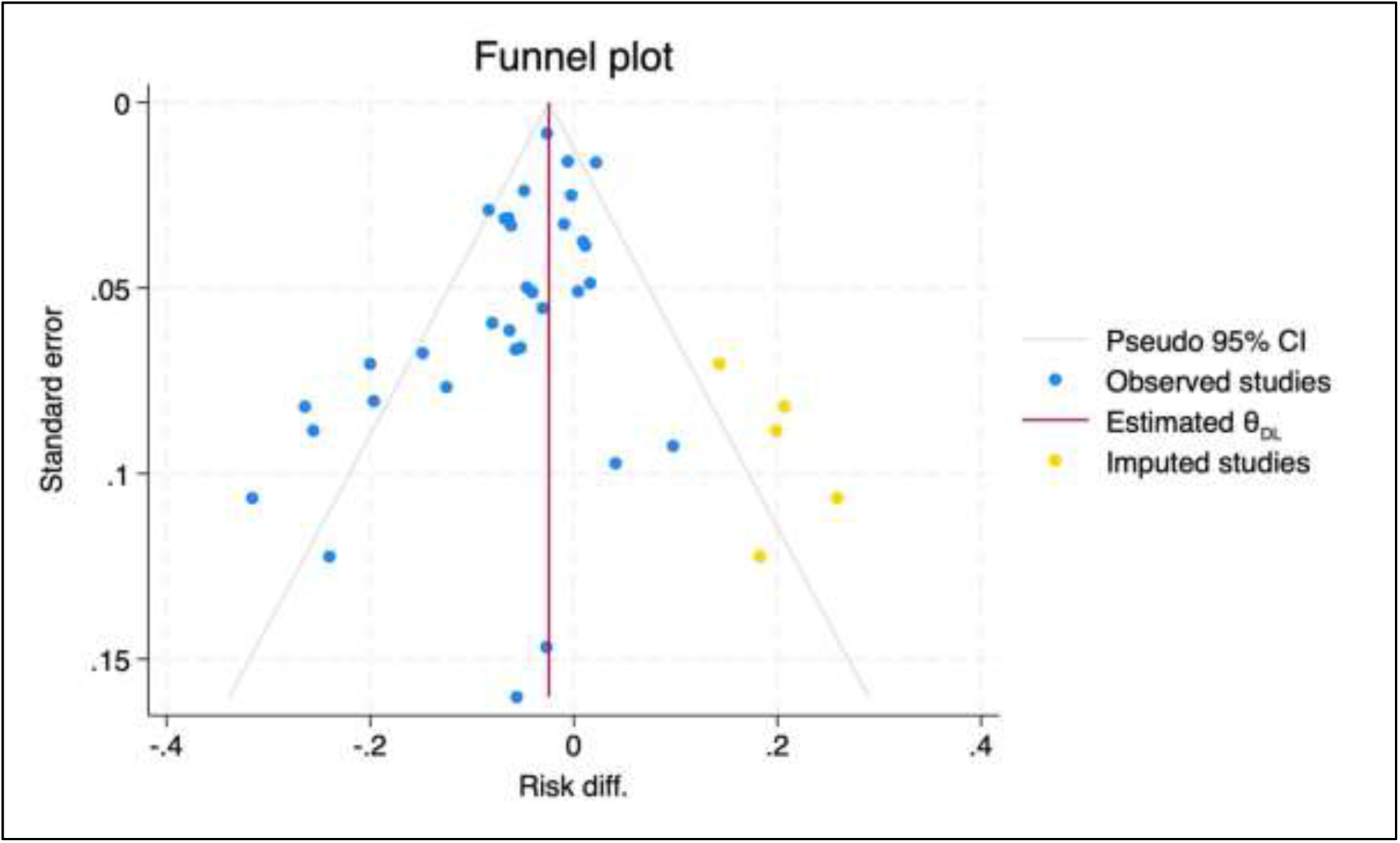

**Figure 7:**
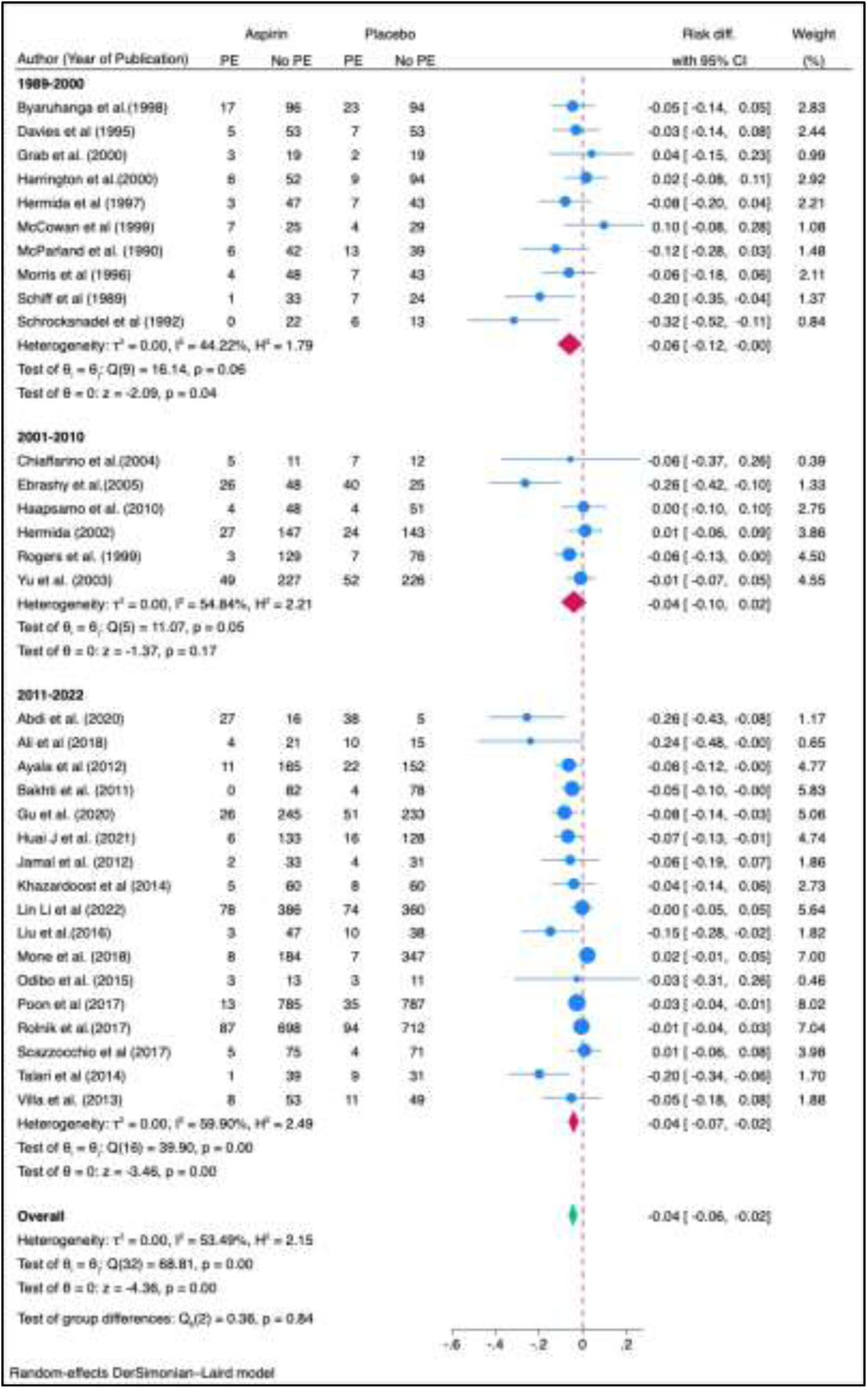
The subgroup analysis of studies investigating the effect of aspirin on PE risk, stratified by year of publication (1989-2022), indicates a significant reduction in PE risk associated with aspirin use. In the 1989-2000 period, the pooled risk difference was −0.06 (95% CI: −0.12 to −0.00), showing a significant reduction in PE risk, with moderate heterogeneity (I^2^ = 44.22%, p = 0.04). For the 2001-2010 period, the pooled risk difference was −0.04 (95% CI: −0.10 to 0.02), but the effect was not statistically significant (p = 0.17), and heterogeneity was higher (I^2^ = 54.84%). In contrast, the 2011-2022 period showed a significant pooled risk difference of −0.04 (95% CI: −0.07 to −0.02), with a notable protective effect of aspirin (p = 0.00), though with higher heterogeneity (I^2^ = 59.90%). The test for group differences was not statistically significant (p = 0.84), suggesting that the effect of aspirin on PE risk did not vary significantly across time periods, although recent studies showed a clearer benefit.

**Figure 8:**
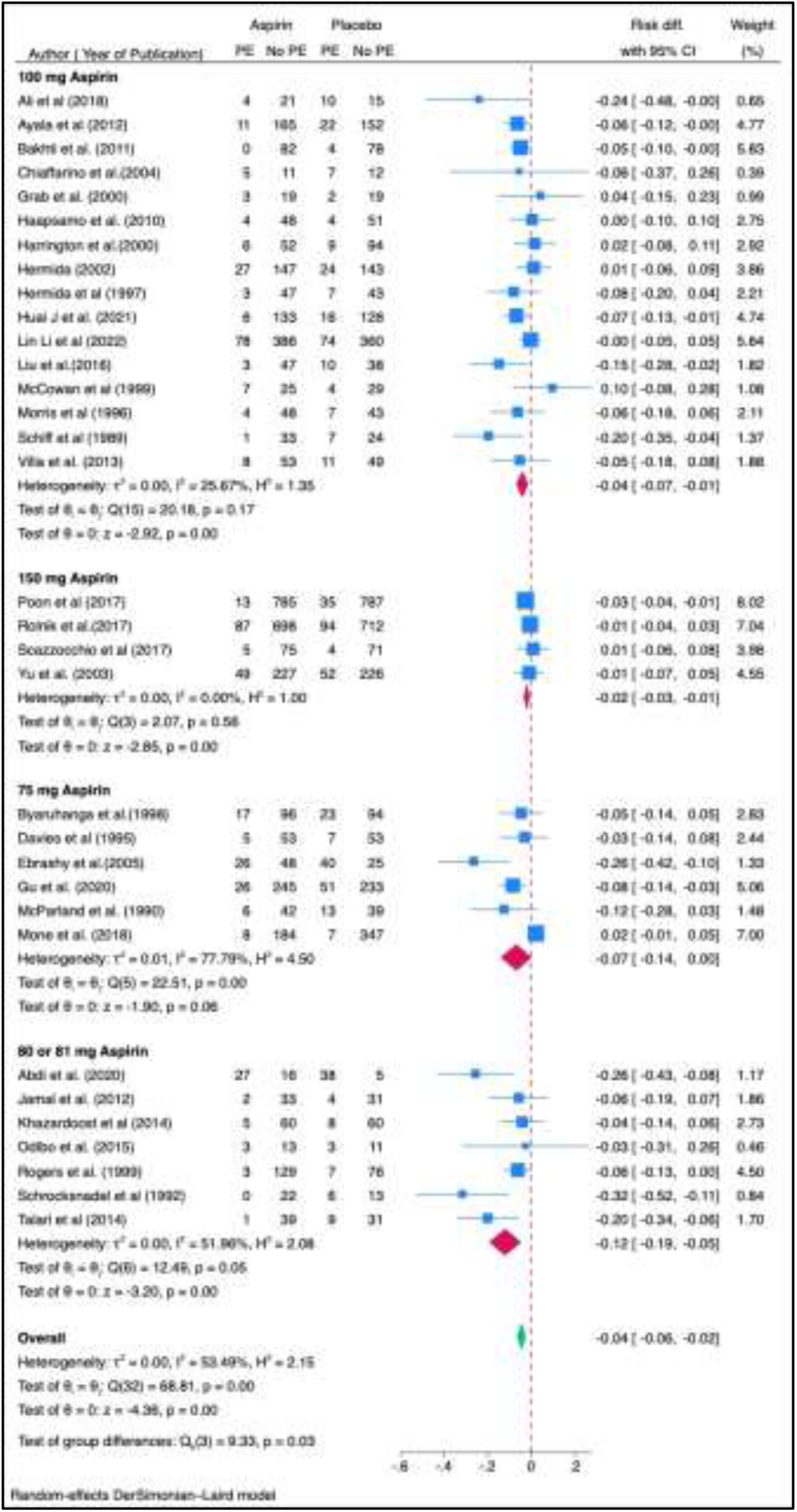
The subgroup analysis based on aspirin dosage reveals different effects on PE risk. For 100 mg aspirin, the pooled risk difference was −0.04 (95% CI: −0.07 to −0.01), showing a significant reduction in PE risk (p = 0.00) with low heterogeneity (I^2^ = 25.67%). For 150 mg aspirin, the pooled risk difference was −0.02 (95% CI: −0.03 to −0.01), also indicating a significant reduction in PE risk (p = 0.00) with no heterogeneity (I^2^ = 0.00%). However, for 75 mg aspirin, the pooled risk difference was −0.07 (95% CI: −0.14 to 0.00), suggesting a non-significant trend toward reduced PE risk (p = 0.06), but with high heterogeneity (I^2^ = 77.79%). For 80 or 81 mg aspirin, the pooled risk difference was −0.12 (95% CI: −0.19 to −0.05), demonstrating a significant protective effect (p = 0.05), with moderate heterogeneity (I^2^ = 51.96%). The test of group differences was significant (p = 0.03), indicating that the protective effect of aspirin on PE risk varied across dosage groups, with 80-81 mg showing the most pronounced effect.

**Figure 9:**
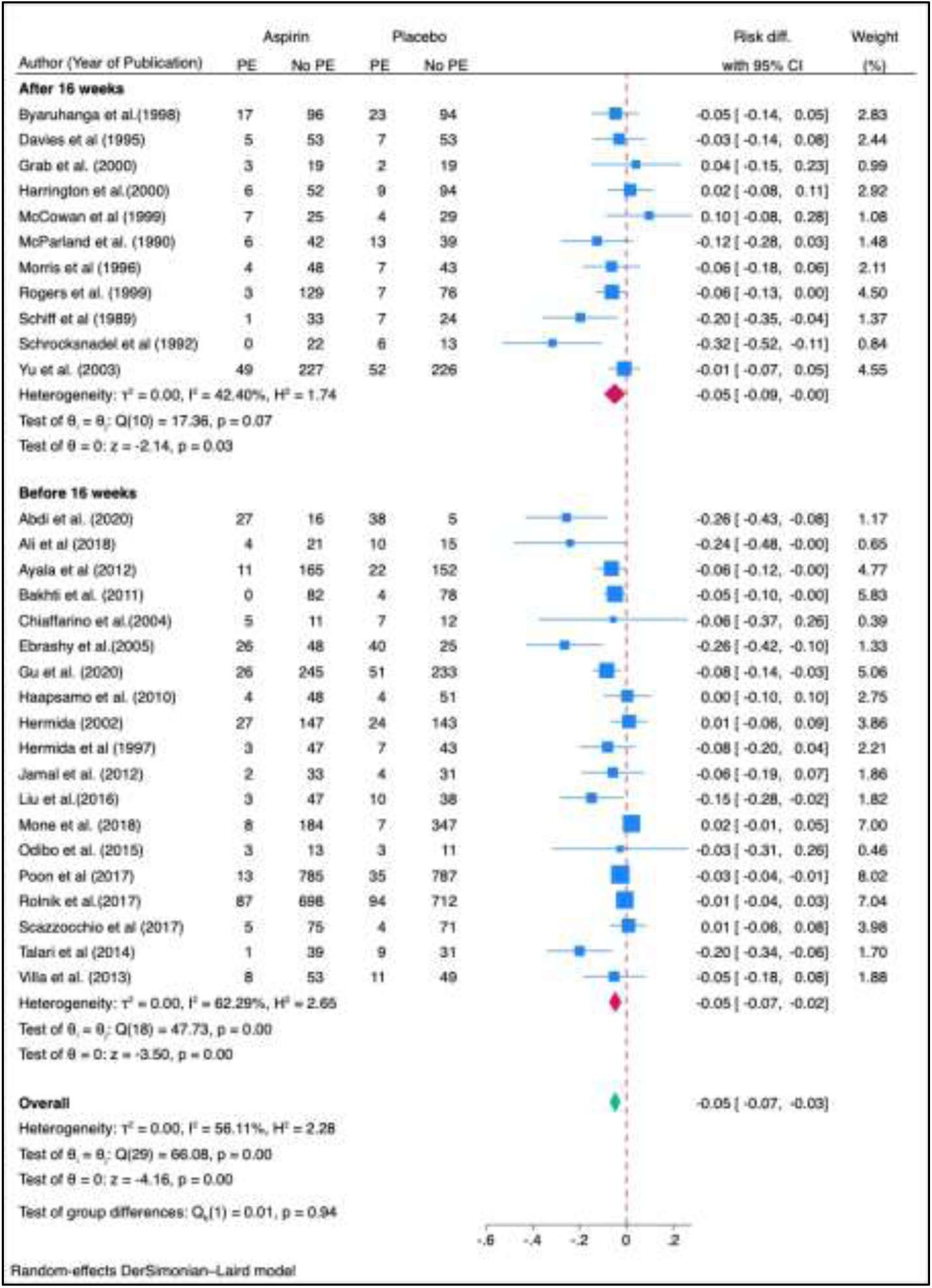
The forest plot illustrates a subgroup analysis of studies examining the effect of aspirin on preventing PE, categorized by the timing of initiation—before or after 16 weeks of gestation. The test for subgroup differences between early and late initiation was not statistically significant (p = 0.94), suggesting that while both strategies reduce PE risk, there is no clear advantage of one timing strategy over the other.

**Figure 10:**
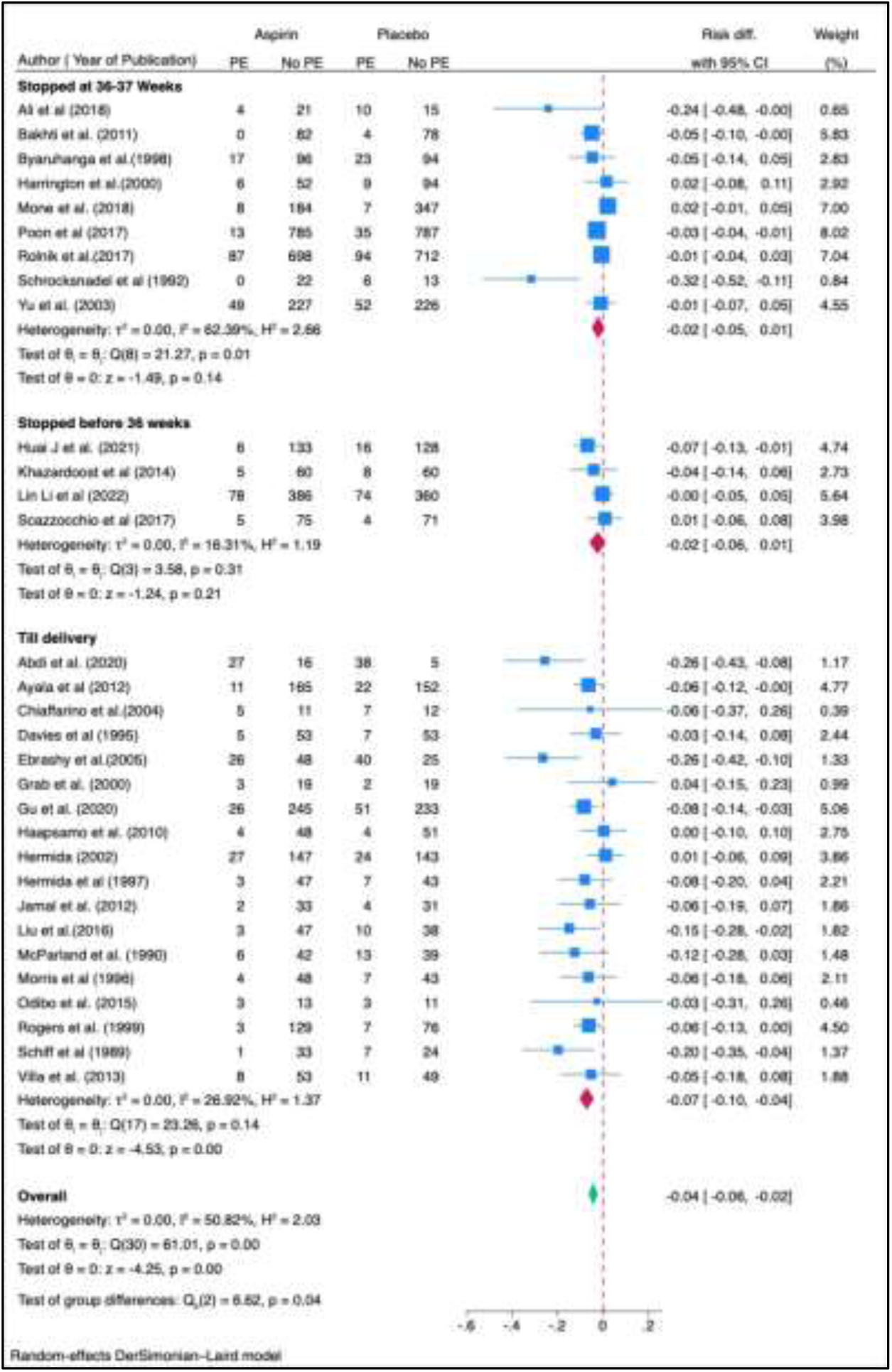
The forest plot displays a subgroup analysis based on the timing of aspirin cessation during pregnancy and its effect on the risk of preeclampsia (PE). The groups are categorized into three: studies where aspirin was stopped at 36–37 weeks, before 36 weeks, and until delivery. The test of group differences (p = 0.04) suggests that the timing of aspirin cessation significantly influences its protective effect against PE. Continuing aspirin until delivery appears to offer the greatest benefit in reducing PE risk.

**Figure 11:**
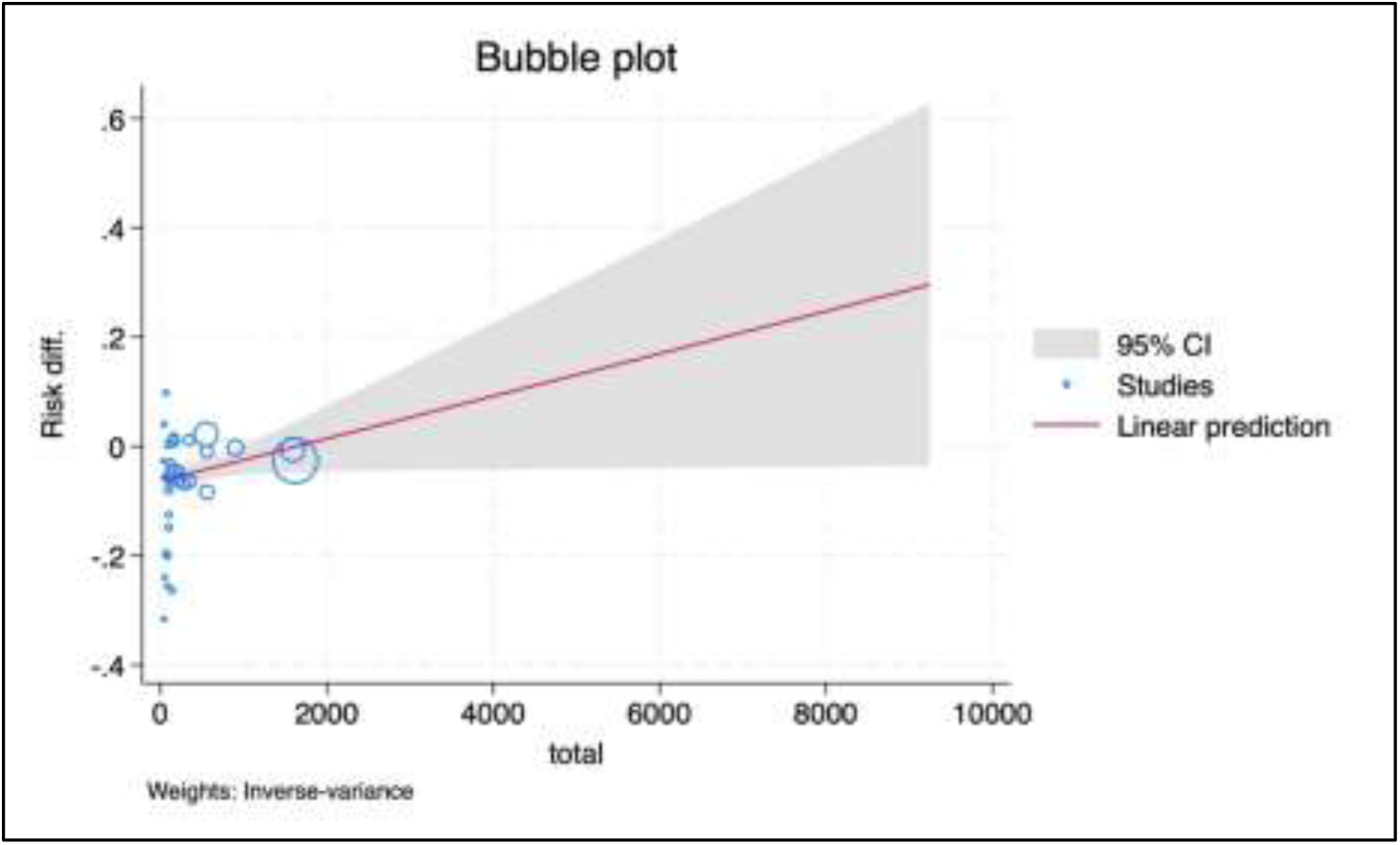
The meta-regression analysis revealed that sample size significantly influences the protective effect of aspirin on PE (Wald chi^2^ = 4.05, p = 0.0441). The positive coefficient indicates that as sample size increases, the protective effect diminishes slightly, with larger studies showing less negative risk differences (i.e., smaller reductions in PE risk) compared to smaller studies, which exhibit greater protective effects (more negative risk differences). Despite this, aspirin’s protective effect remains significant across all studies.

**Figure 12:**
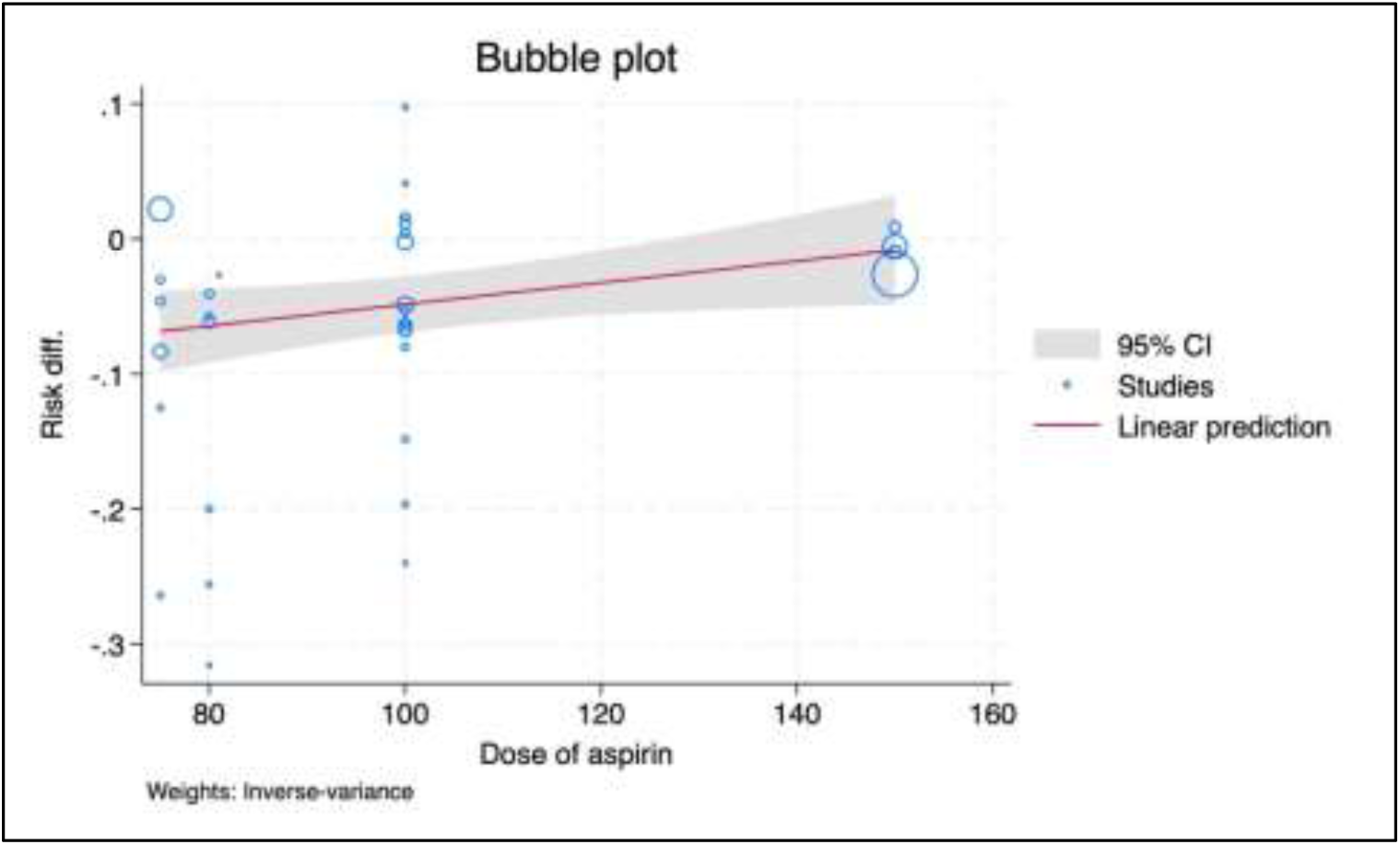
The meta-regression analysis examining the influence of aspirin dose on the risk difference for PE revealed a significant effect (Wald chi^2^ = 4.34, p = 0.0372). The positive coefficient suggests that as the dose of aspirin increases, the protective effect against PE diminishes slightly, with higher doses associated with less negative risk differences. Conversely, lower doses of aspirin showed greater protective effects.

**Figure 13:**
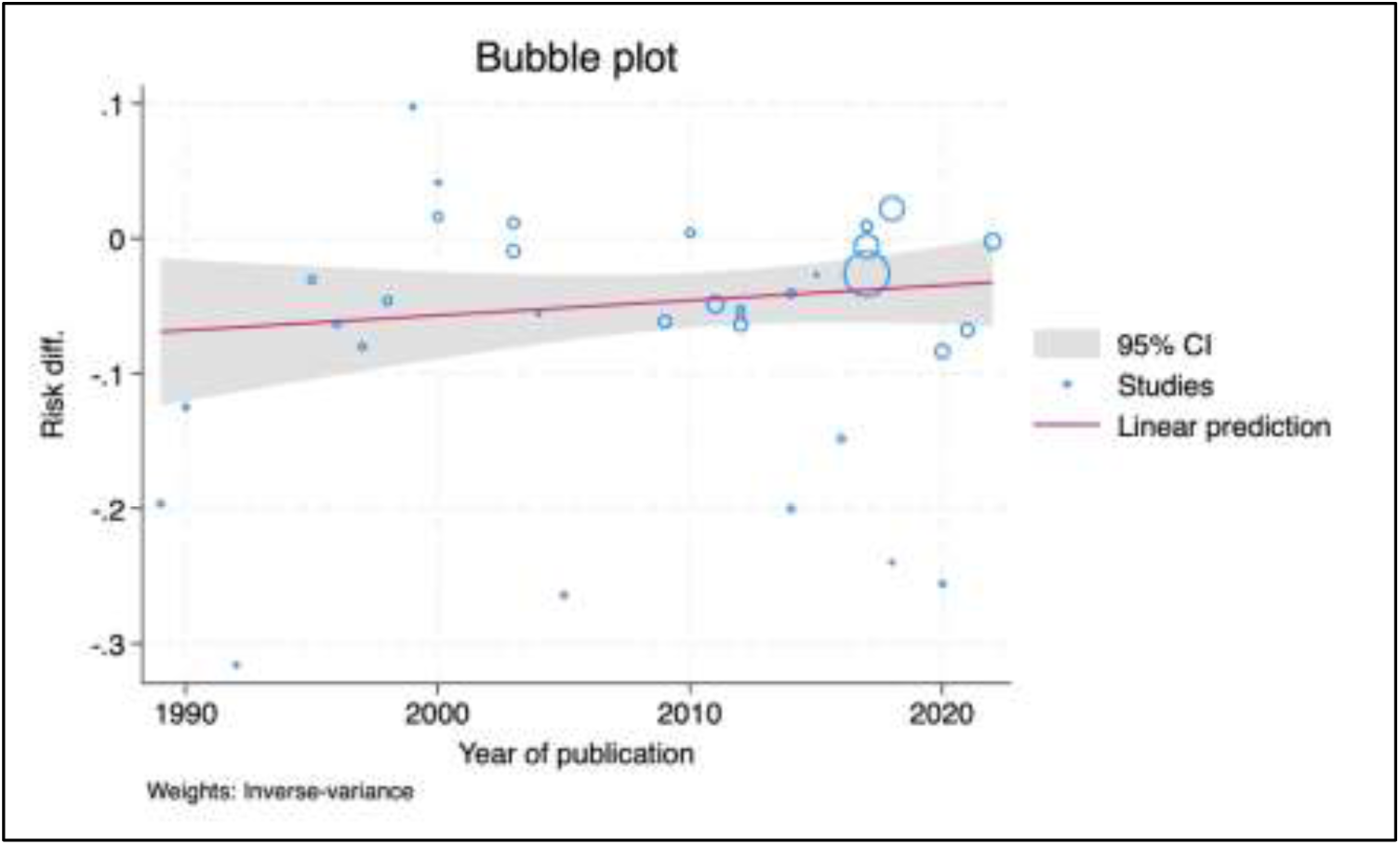
The meta-regression analysis examining the year of publication as a predictor of risk difference for preeclampsia (PE) showed no significant effect (Wald chi^2^ = 0.90, p = 0.3424

**Figure 14:**
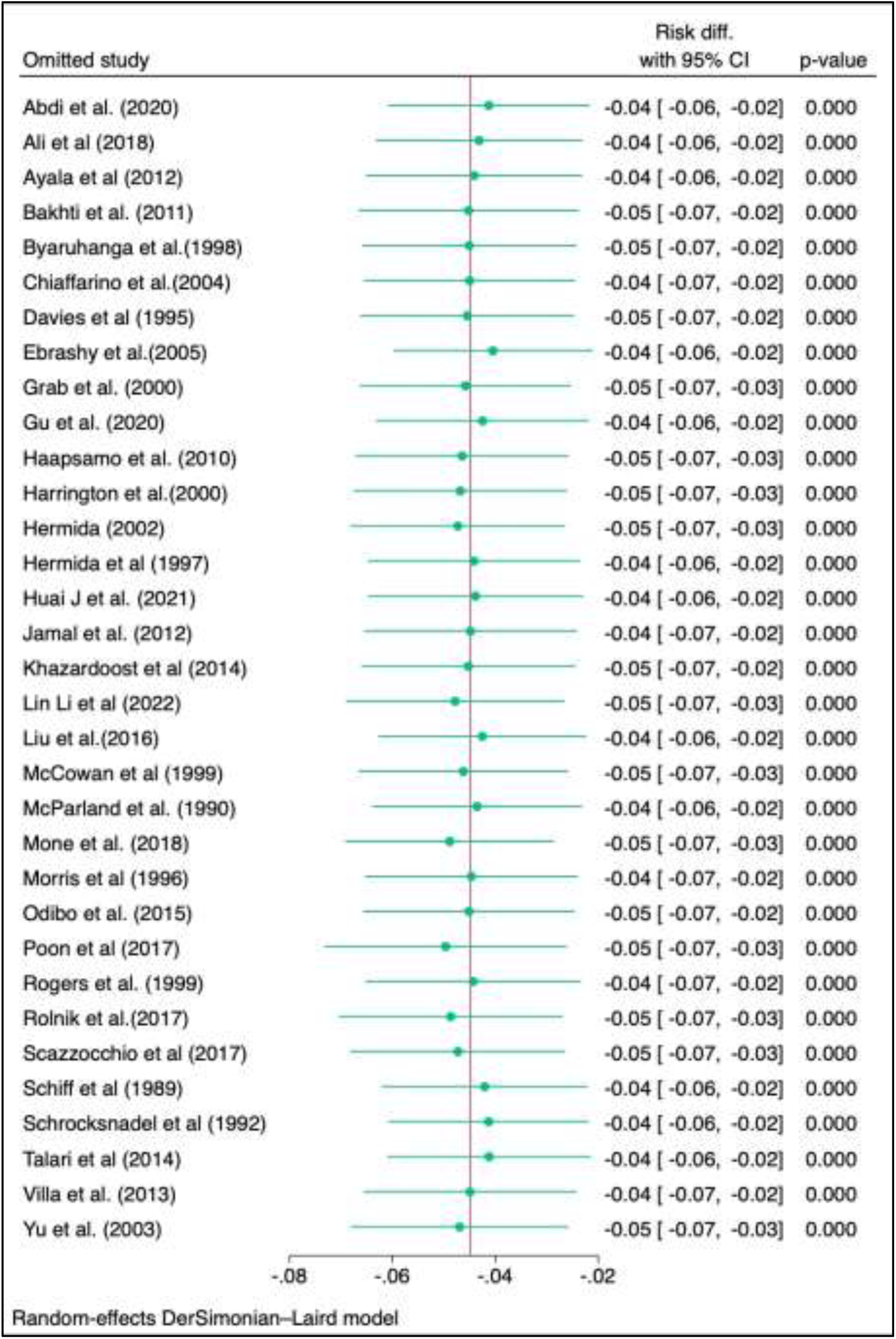
The “leave-one-out” sensitivity analysis suggests that the overall risk difference is stable, regardless of which study is omitted. When each individual study is removed, the risk difference remains consistent at approximately −0.04 to −0.05, with narrow confidence intervals that do not cross zero, and all p-values remain significant (p < 0.05). This indicates that no single study unduly influences the overall result, suggesting the findings are robust

**Figure 15:**
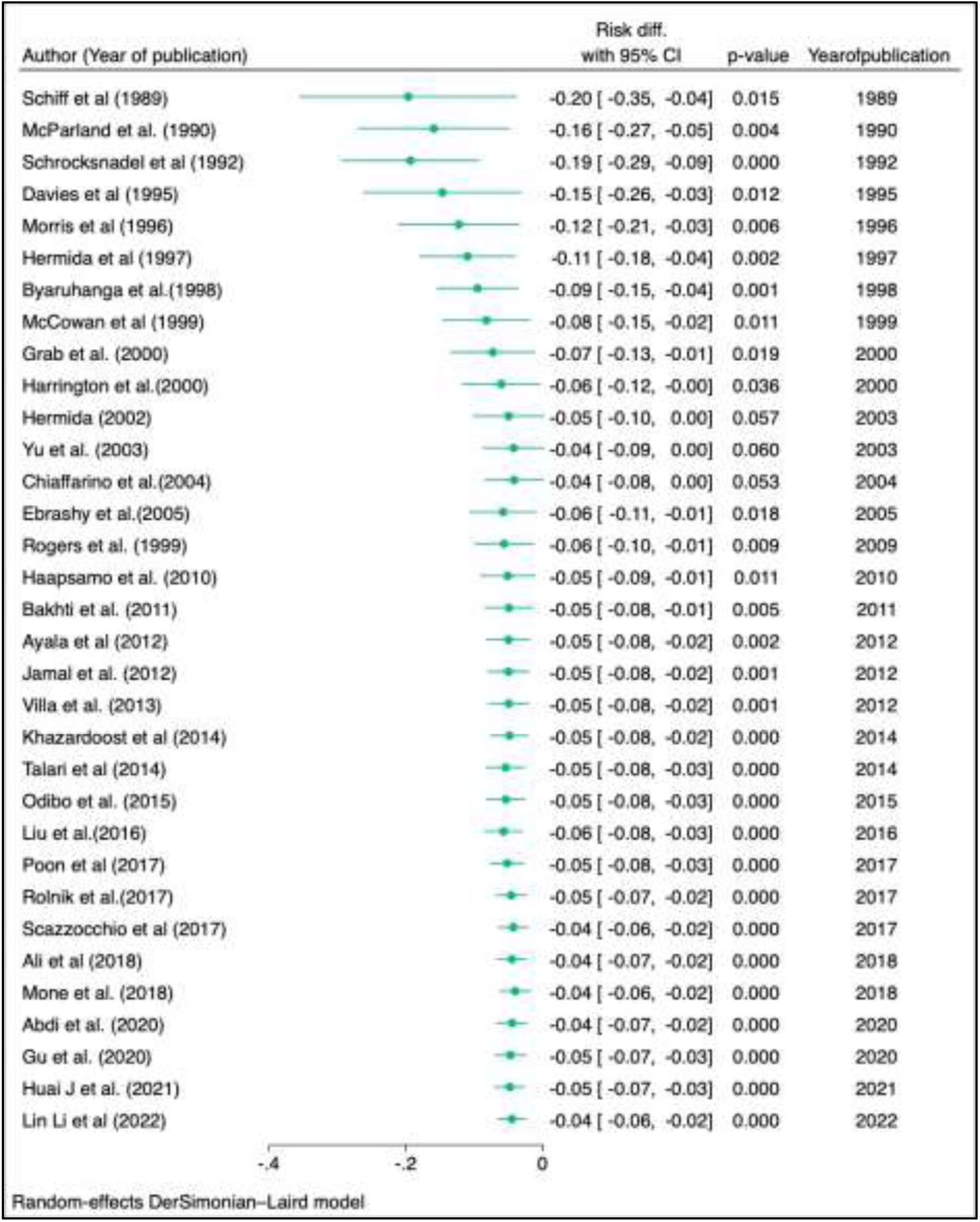
The cumulative meta-analysis presented in the graph demonstrates how the effect size of aspirin’s risk difference on preeclampsia has evolved as additional studies were published over time. The plot begins with studies from 1989, where the risk difference for preeclampsia was significantly reduced in aspirin-treated groups compared to placebo. As more studies were added, the cumulative effect size stabilized around −0.04 by 2022. Each study added precision to the estimate, reducing the confidence intervals over time, especially from the mid-2000s onward. All cumulative estimates are statistically significant, consistently showing a negative risk difference, indicating a beneficial effect of aspirin in reducing preeclampsia risk. Overall, this plot underscores the robustness and stability of the aspirin effect as evidence has accumulated, with the final cumulative effect size showing a significant reduction in preeclampsia risk by approximately 4%.

**Figure 16:**
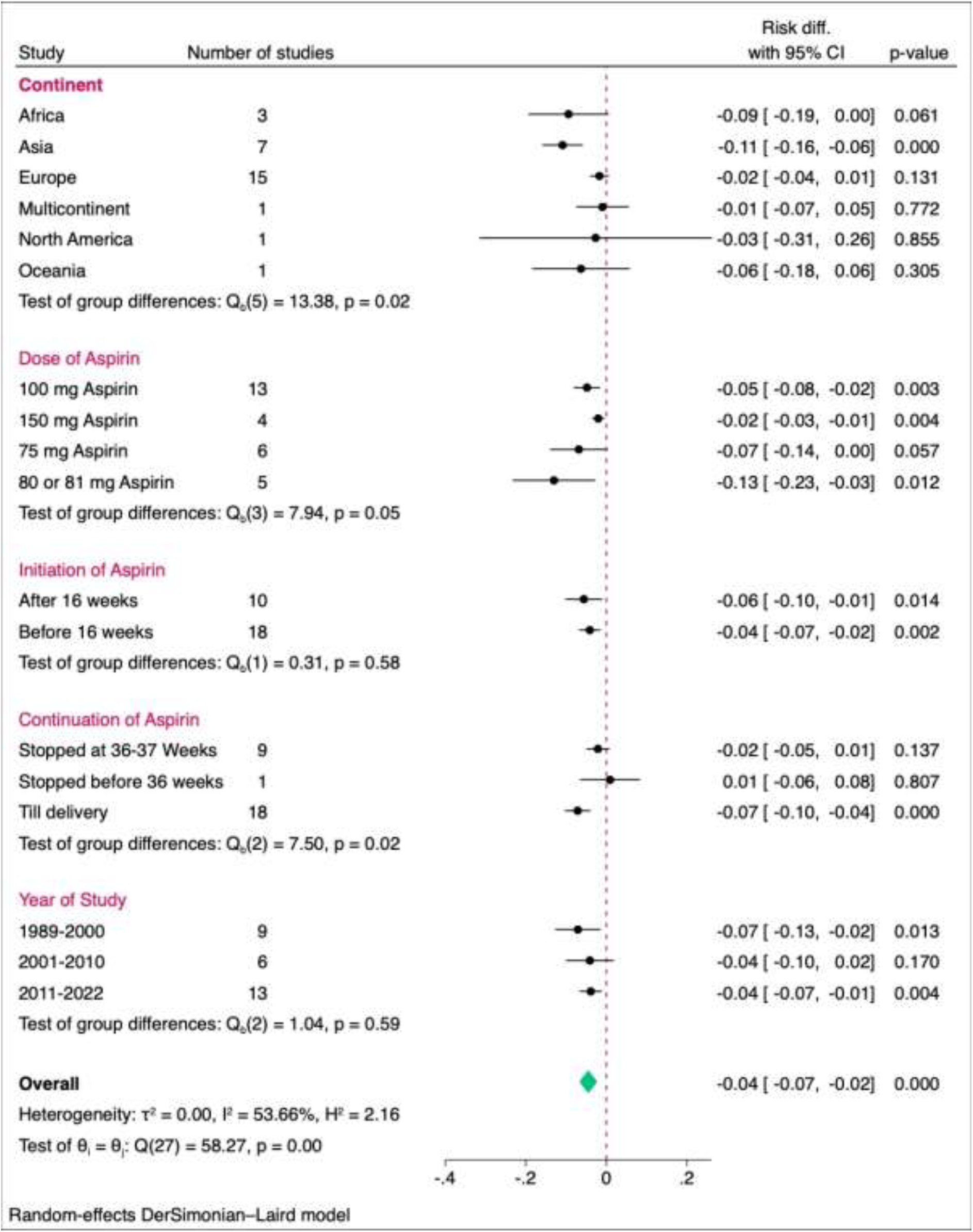

## Data Availability

All data produced in the present study are available upon reasonable request to the authors

## Participation of each author

Conceptualization: Dr. Upma Saxena, Dr. Abhishek Lachyan

Data curation: Dr. Abhishek Lachyan, Dr. Aninda Debnath, Dr. Sunanda Gupta, Dr. Ankit Yadav

Formal Analysis: Dr. Abhishek Lachyan, Dr. Aninda Debnath, Dr. Sunanda Gupta, Dr. Ankit Yadav, Dr. Rishi Gupta

Funding acquisition: ICMR

Investigation: Dr. Upma Saxena, Dr. Abhishek Lachyan

Methodology: Dr. Upma Saxena, Dr. Abhishek Lachyan, Dr. Aninda Debnath

Project administration: Dr. Upma Saxena, Dr. Abhishek Lachyan

Resources: Dr. Jugal Kishore, Dr. Rishi Gupta, Dr. Sidarrth Prasad, Dr. Chanchal Goyal, Dr. Anju Sinha

Software: SPSS, Stata

Supervision: Dr. Upma Saxena, Dr. Abhishek Lachyan

Validation: Dr. Upma Saxena, Dr. Abhishek Lachyan, Dr. Aninda Debnath, Dr. Sunanda Gupta, Ankit Yadav, Dr. Jugal Kishore, Dr. Rishi Gupta, Dr. Sidarrth Prasad, Dr. Chanchal Goyal, Dr. Anju Sinha.

## ACKNOWLEDGEMENT

I appreciate the encouragement from my friends, colleagues, and family. Lastly, I thank all the patients who participated in this study.

We would like to acknowledge the Indian Council of Medical Research (ICMR) for funding support, which facilitated the initiation of this project.

## Meta-analysis and Meta-regression on the Effect of Aspirin vs Placebo and High Dose vs Low Dose on Preeclampsia

### Introduction

This meta-analysis investigates the effect of aspirin versus placebo on the reduction of preeclampsia (PE) and whether there is a dose-response effect (high dose versus low dose). We perform two key analyses:

1. A comparison between aspirin and placebo to evaluate their impact on reducing preeclampsia.
2. A meta-regression to assess whether high-dose or low-dose aspirin leads to a greater reduction in preeclampsia.

### Meta-analysis Results: Aspirin vs Placebo

The forest plot below presents the overall effect of aspirin versus placebo on preeclampsia, with individual studies contributing to the overall pooled estimate.

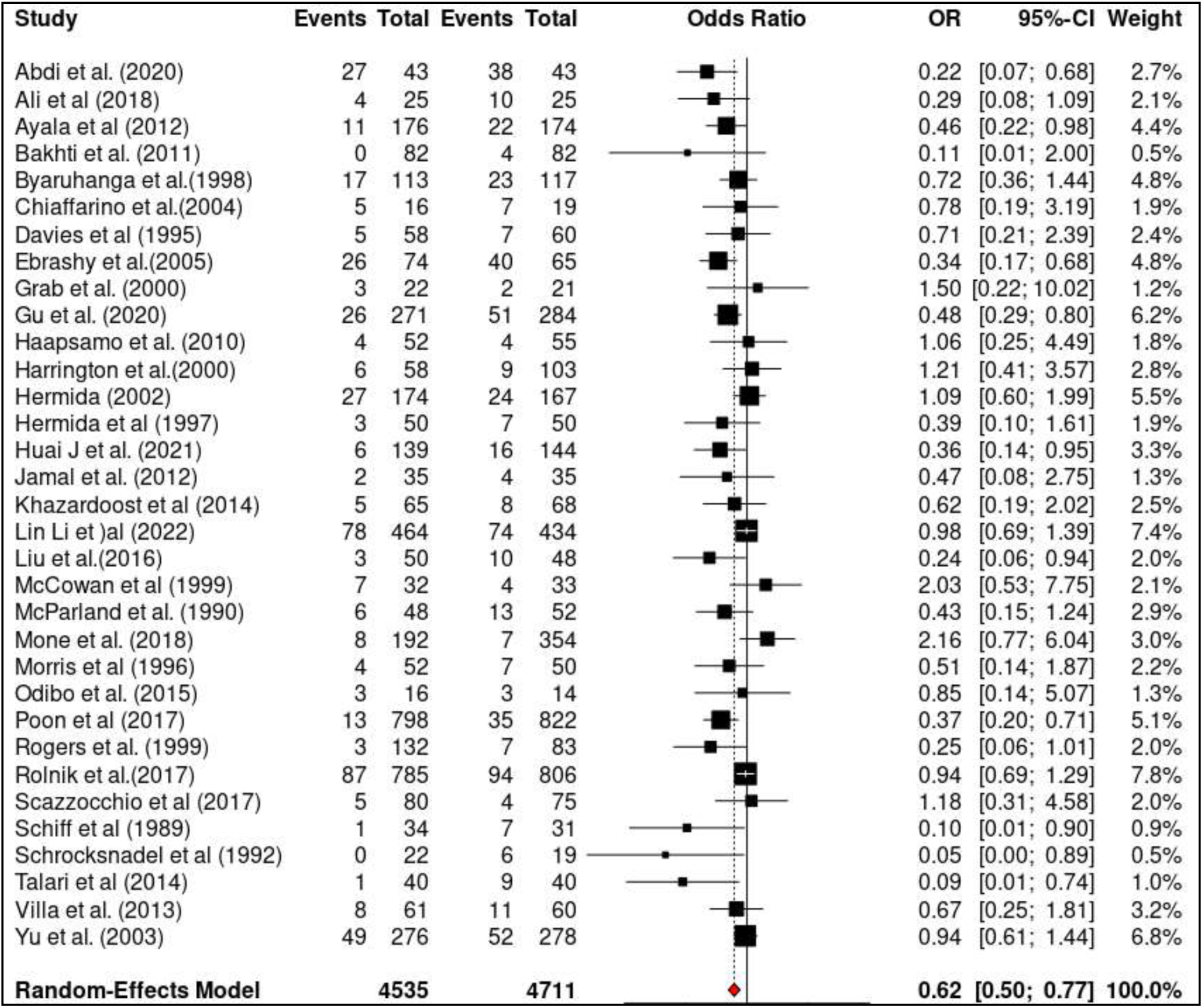

#### Forest plot of aspirin vs placebo effect on preeclampsia

The meta-analysis included 33 studies, with a total of 4535 participants in the aspirin group and 4711 in the placebo group. The overall odds ratio from the random-effects model is 0.62 (95% CI: 0.5 to 0.77, p = <0.001). This indicates that aspirin significantly reduces the risk of preeclampsia compared to placebo.

The I^2^ statistic for heterogeneity is 0.4%, indicating low to moderate heterogeneity across studies (Q = 57.24, p = 0.00398). The significant Q-test result suggests the presence of heterogeneity, warranting further exploration of potential moderating factors.

### Meta-regression Results: High Dose vs Low Dose

The meta-regression assesses the dose-response effect between high-dose and low-dose aspirin on preeclampsia reduction. The bubble plot below visualizes this relationship.

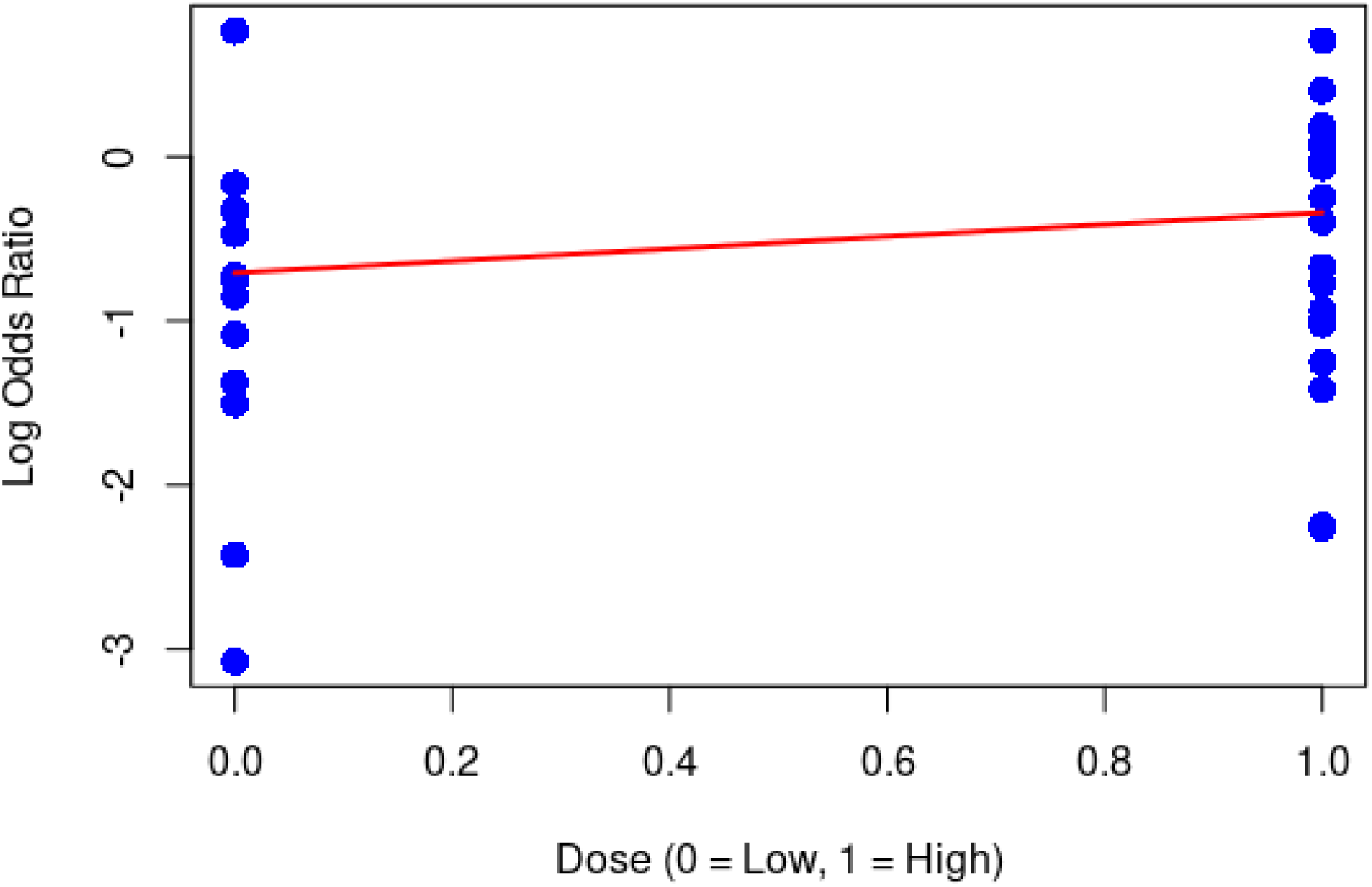

#### Meta-regression bubble plot of dose effect on preeclampsia reduction

The meta-regression coefficient for dose is 0.37 (95% CI: −0.06 to 0.8, p = 0.090). The positive coefficient suggests that high-dose aspirin may be more effective than low-dose in reducing preeclampsia. However, the confidence interval includes zero, and the p-value is above the conventional 0.05 significance level, indicating that the dose-response effect is not statistically significant.

The meta-regression model explains 28.04% of the heterogeneity in effect sizes (Q_M = 2.87, p = 0.0902).

### Conclusion

In this meta-analysis of 33 studies with a total of 9246 participants, we found that aspirin significantly reduces the risk of preeclampsia compared to placebo (OR = 0.62, 95% CI: 0.5 to 0.77, p = <0.001).

The meta-regression analysis suggests a potential dose-response effect, with high-dose aspirin possibly leading to a greater reduction in preeclampsia compared to low-dose aspirin (β = 0.37, 95% CI: −0.06 to 0.8, p = 0.090). However, this effect is not statistically significant, and further research is needed to confirm the dose-response relationship.

The low to moderate heterogeneity observed in the meta-analysis (I^2^ = 0.4%, Q = 57.24, p = 0.004) suggests the presence of potential moderating factors that may influence the treatment effect. Further investigation into these factors is warranted to better understand the variability in the results.

These findings provide evidence for the use of aspirin in the prevention of preeclampsia, with a significant reduction in risk compared to placebo. However, the lack of a statistically significant dose-response effect and the presence of heterogeneity highlight the need for further research to confirm the optimal dose and identify potential moderating factors.

**Annexure 1:**
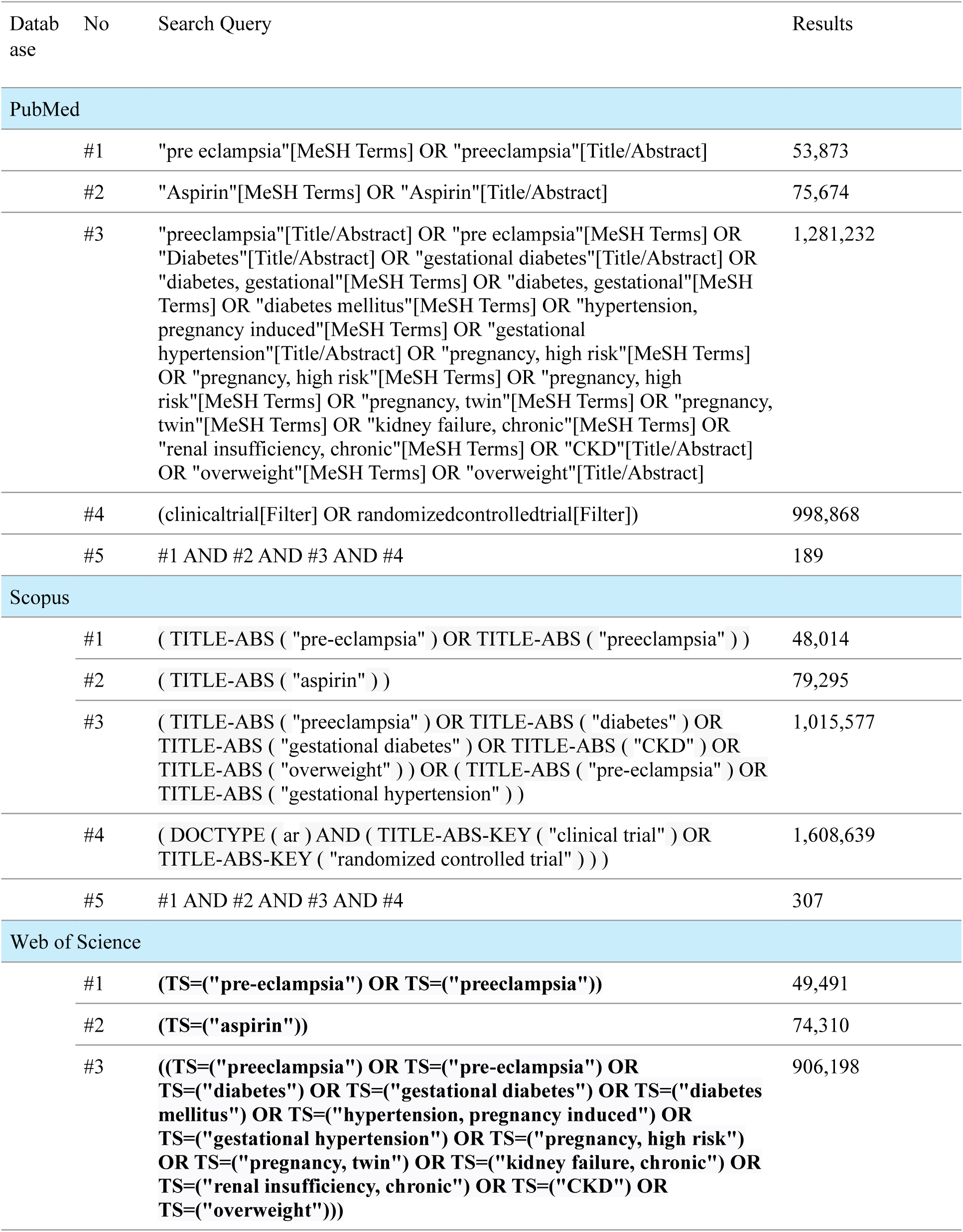
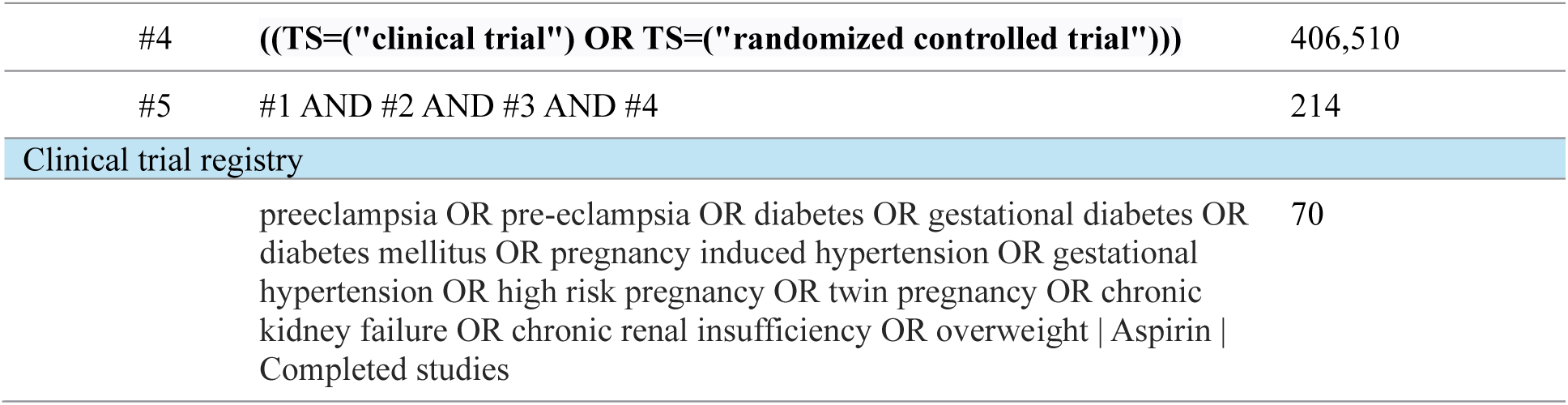
Search strategy (31/07/24)

## References

1. Ives, C, Sinkey, R, Rajapreyar, I. et al. Preeclampsia—Pathophysiology and Clinical Presentations: JACC State-of-the-Art Review. JACC. 2020 Oct, 76 (14) 1690–1702. a. 10.1016/j.jacc.2020.08.014.

2. Nelson KM, Irvin-Choy N, Hoffman MK, Gleghorn JP, Day ES. Diseases and conditions that impact maternal and fetal health and the potential for nanomedicine therapies. Adv Drug Deliv Rev. 2021 Mar;170:425–438. doi: 10.1016/j.addr.2020.09.013. Epub 2020 Sep 28. PMID: 33002575; PMCID: PMC7981251.

3. Xiao Y, Ling Q, Yao M, Gu Y, Lan Y, Liu S, Yin J, Ma Q. Aspirin 75 mg to prevent preeclampsia in high-risk pregnancies: a retrospective real-world study in China. Eur J Med Res. 2023 Feb 2;28(1):56. doi: 10.1186/s40001-023-01024-7. PMID: 36732824; PMCID: PMC9893656.

4. Atallah A, Lecarpentier E, Goffinet F, Doret-Dion M, Gaucherand P, Tsatsaris V. Aspirin for Prevention of Preeclampsia. Drugs. 2017 Nov;77(17):1819–1831. doi: 10.1007/s40265-017-0823-0. PMID: 29039130; PMCID: PMC5681618.

5. Lin L, Huai J, Li B, Zhu Y, Juan J, Zhang M, Cui S, Zhao X, Ma Y, Zhao Y, Mi Y, Ding H, Chen D, Zhang W, Qi H, Li X, Li G, Chen J, Zhang H, Yu M, Sun X, Yang H. A randomized controlled trial of low-dose aspirin for the prevention of preeclampsia in women at high risk in China. Am J Obstet Gynecol. 2022 Feb;226(2):251.e1-251.e12. doi: 10.1016/j.ajog.2021.08.004. Epub 2021 Aug 10. PMID: 34389292.

6. Van Doorn R, Mukhtarova N, Flyke IP, Lasarev M, Kim K, Hennekens CH, Hoppe KK. Dose of aspirin to prevent preterm preeclampsia in women with moderate or high-risk factors: A systematic review and meta-analysis. PLoS One. 2021 Mar 9;16(3):e0247782. doi: 10.1371/journal.pone.0247782. PMID: 33690642; PMCID: PMC7943022.

7. Abdi N, Rozrokh A, Alavi A, Zare S, Vafaei H, Asadi N, Kasraeian M, Hessami K. The effect of aspirin on preeclampsia, intrauterine growth restriction and preterm delivery among healthy pregnancies with a history of preeclampsia. J Chin Med Assoc. 2020 Sep;83(9):852–857. doi: 10.1097/JCMA.0000000000000400. PMID: 32773581; PMCID: PMC7478204.

8. Jalal Ali, P. and Najmaddin Fattah, C. (2018) “Effect of Low Dose Aspirin in Low Risk Pregnant Ladies with Abnormal Uterine Artery Doppler Results, and the Evaluation of Maternal and Fetal outcomes: A Randomized Clinical Trial”, Kurdistan Journal of Applied Research, 3(2), pp. 85–89. doi:10.24017/science.2018.2.14.

9. Ayala DE, Ucieda R, Hermida RC. Chronotherapy with low-dose aspirin for prevention of complications in pregnancy. Chronobiol Int. 2013;30(1-2):260–279. doi:10.3109/07420528.2012.717455.

10. Bakhti A, Vaiman D. Prevention of gravidic endothelial hypertension by aspirin treatment administered from the 8th week of gestation [published correction appears in Hypertens Res. 2012 Feb;35(2):244]. Hypertens Res. 2011;34(10):1116–1120. doi:10.1038/hr.2011.111.

11. Byaruhanga RN, Chipato T, Rusakaniko S. A randomized controlled trial of low-dose aspirin in women at risk from pre-eclampsia. Int J Gynaecol Obstet. 1998 Feb;60(2):129–35. doi: 10.1016/s0020-7292(97)00257-9. PMID: 9509950.

12. Chiaffarino F, Parazzini F, Paladini D, Acaia B, Ossola W, Marozio L, et al. A small randomised trial of low-dose aspirin in women at high risk of pre-eclampsia. European Journal of Obstetrics and Gynecology and Reproductive Biology. 2004 Feb 10;112(2):142–4.

13. Davies NJ, Gazvani MR, Farquharson RG, Walkinshaw SA. Low-Dose Aspirin in the Prevention of Hypertensive Disorders of Pregnancy in Relatively Low-Risk Nulliparous Women. Hypertension in Pregnancy. 1995 Jan;14(1):49–55.

14. Ebrashy A, Ibrahim M, Marzook A, Yousef D. Usefulness of aspirin therapy in high-risk pregnant women with abnormal uterine artery Doppler ultrasound at 14-16 weeks pregnancy: randomized controlled clinical trial. Croat Med J. 2005 Oct;46(5):826–31. PMID: 16158479.

15. Grab D, Paulus WE, Erdmann M, et al. Effects of low-dose aspirin on uterine and fetal blood flow during pregnancy: results of a randomized, placebo-controlled, double-blind trial. Ultrasound Obstet Gynecol. 2000;15(1):19–27. doi:10.1046/j.1469-0705.2000.00009.x

16. Gu W, Lin J, Hou YY, Lin N, Song MF, Zeng WJ, et al. Effects of low-dose aspirin on the prevention of preeclampsia and pregnancy outcomes: A randomized controlled trial from Shanghai, China. Eur J Obstet Gynecol Reprod Biol. 2020 May;248:156–63.

17. Haapsamo M, Martikainen H, Tinkanen H, Heinonen S, Nuojua-Huttunen S, Räsänen J. Low-dose aspirin therapy and hypertensive pregnancy complications in unselected IVF and ICSI patients: a randomized, placebo-controlled, double-blind study. Hum Reprod. 2010;25(12):2972–2977. doi:10.1093/humrep/deq286

18. Harrington K, Kurdi W, Aquilina J, England P, Campbell S. A prospective management study of slow-release aspirin in the palliation of uteroplacental insufficiency predicted by uterine artery Doppler at 20 weeks. Ultrasound Obstet Gynecol. 2000 Jan;15(1):13-doi: 10.1046/j.1469-0705.2000.00002.x. PMID: 10776007.

19. Hermida RC, Ayala DE, Iglesias M. Administration time-dependent influence of aspirin on blood pressure in pregnant women. Hypertension. 2003;41(3 Pt 2):651–656. doi: 10.1161/01.HYP.0000047876.63997.EE

20. Hermida RC, Ayala DE, Iglesias M, et al. Time-dependent effects of low-dose aspirin administration on blood pressure in pregnant women. Hypertension. 1997;30(3 Pt 2):589–595. doi:10.1161/01.hyp.30.3.589.

21. Huai J, Lin L, Juan J, Chen J, Li B, Zhu Y, Yu M, Yang H. Preventive effect of aspirin on preeclampsia in high-risk pregnant women with stage 1 hypertension. J Clin Hypertens (Greenwich). 2021 May;23(5):1060–1067. doi: 10.1111/jch.14149. Epub 2021 Jan 5. PMID: 33400389; PMCID: PMC8678830.

22. Jamal A, Milani F, Al-Yasin A. Evaluation of the effect of metformin and aspirin on utero placental circulation of pregnant women with PCOS. Iran J Reprod Med. 2012;10(3):265–270.

23. Khazardoost S, Mousavi S, Borna S, Hantoushzadeh S, Alavi A, Khezerlou N. Effect of aspirin in prevention of adverse pregnancy outcome in women with elevated alpha-fetoprotein. J Matern Fetal Neonatal Med. 2014;27(6):561–565. doi:10.3109/14767058.2013.822483

24. Xiaoyan Lin, Jingchao Yong, Ming Gan, Shaowen Tang & Jiangbo Du (2024) Impact of low-dose aspirin exposure on obstetrical outcomes: a meta-analysis, Journal of Psychosomatic Obstetrics & Gynecology, 45:1, 2344079, DOI: 10.1080/0167482X.2024.2344079.

25. Liu F, Yang H, Li G, Zou K, Chen Y. Effect of a small dose of aspirin on quantitative test of 24-h urinary protein in patients with hypertension in pregnancy. Exp Ther Med. 2017 Jan;13(1):37–40. doi: 10.3892/etm.2016.3924. Epub 2016 Nov 22. PMID: 28123464; PMCID: PMC5244777.

26. McCowan LM, Harding J, Roberts A, Barker S, Ford C, Stewart A. Administration of low-dose aspirin to mothers with small for gestational age fetuses and abnormal umbilical Doppler studies to increase birthweight: a randomised double-blind controlled trial. Br J Obstet Gynaecol. 1999;106(7):647–651. doi:10.1111/j.1471-0528.1999.tb08362.x

27. McParland P, Pearce JM, Chamberlain GVP. Doppler ultrasound and aspirin in recognition and prevention of pregnancy-induced hypertension. The Lancet. 1990 Jun 30;335(8705):1552–5.

28. Mone F, Mulcahy C, McParland P, et al. Trial of feasibility and acceptability of routine low-dose aspirin versus Early Screening Test indicated aspirin for pre-eclampsia prevention (TEST study): a multicentre randomised controlled trial. BMJ Open 2018;8:e022056. doi:10.1136/bmjopen-2018-022056.

29. Morris JM, Fay RA, Ellwood DA, Cook CM, Devonald KJ. A randomized controlled trial of aspirin in patients with abnormal uterine artery blood flow. Obstet Gynecol. 1996 Jan;87(1):74–8. doi: 10.1016/0029-7844(95)00340-1. PMID: 8532271.

30. Odibo AO, Goetzinger KR, Odibo L, Tuuli MG. Early prediction and aspirin for prevention of pre-eclampsia (EPAPP) study: a randomized controlled trial. Ultrasound Obstet Gynecol. 2015 Oct;46(4):414–8. doi: 10.1002/uog.14889. Epub 2015 Aug 31. PMID: 25914193.

31. Poon LC, Wright D, Rolnik DL, et al. Aspirin for Evidence-Based Preeclampsia Prevention trial: effect of aspirin in prevention of preterm preeclampsia in subgroups of women according to their characteristics and medical and obstetrical history [published correction appears in Am J Obstet Gynecol. 2018 Apr;218(4):454. doi: 10.1016/j.ajog.2018.01.002]. Am J Obstet Gynecol. 2017;217(5):585.e1–585.e5. doi:10.1016/j.ajog.2017.07.038

32. Rogers MS, Fung HY, Hung CY. Calcium and low-dose aspirin prophylaxis in women at high risk of pregnancy-induced hypertension. Hypertens Pregnancy. 1999;18(2):165–72. doi: 10.3109/10641959909023076. PMID: 10476618.

33. Rolnik DL, Wright D, Poon LCY, et al. ASPRE trial: performance of screening for preterm pre-eclampsia [published correction appears in Ultrasound Obstet Gynecol. 2017 Dec;50(6):807. doi: 10.1002/uog.18950]. Ultrasound Obstet Gynecol. 2017;50(4):492–495. doi:10.1002/uog.18816.

34. Scazzocchio E, Oros D, Diaz D, Ramirez JC, Ricart M, Meler E, González de Agüero R, Gratacos E, Figueras F. Impact of aspirin on trophoblastic invasion in women with abnormal uterine artery Doppler at 11-14 weeks: a randomized controlled study. Ultrasound Obstet Gynecol. 2017 Apr;49(4):435–441. doi: 10.1002/uog.17351. PMID: 27807890.

35. Schiff E, Peleg E, Goldenberg M, Rosenthal T, Ruppin E, Tamarkin M, Barkai G, Ben-Baruch G, Yahal I, Blankstein J, et al. The use of aspirin to prevent pregnancy-induced hypertension and lower the ratio of thromboxane A2 to prostacyclin in relatively high risk pregnancies. N Engl J Med. 1989 Aug 10;321(6):351–6. doi: 10.1056/NEJM198908103210603. PMID: 2664522.

36. Schröcksnadel H, Sitte B, Alge A, Steckel-Berger G, Schwegel P, Pastner E, Daxenbichler G, Hansen H, Dapunt O. Low-dose aspirin in primigravidae with positive roll-over test. Gynecol Obstet Invest. 1992;34(3):146–50. doi: 10.1159/000292748. PMID: 1427414.

37. Talari H, Mesdaghinia E, Abedzadeh Kalahroudi M. Aspirin and preeclampsia prevention in patients with abnormal uterine artery blood flow. Iran Red Crescent Med J. 2014 Aug;16(8):e17175. doi: 10.5812/ircmj.17175. Epub 2014 Aug 5. PMID: 25389483; PMCID: PMC4222009.

38. Villa PM, Kajantie E, Räikkönen K, Pesonen AK, Hämäläinen E, Vainio M, Taipale P, Laivuori H; PREDO Study group. Aspirin in the prevention of pre-eclampsia in high-risk women: a randomised placebo-controlled PREDO Trial and a meta-analysis of randomised trials. BJOG. 2013 Jan;120(1):64–74. doi: 10.1111/j.1471-0528.2012.03493.x. Epub 2012 Nov 6. PMID: 23126307.

39. Yu CK, Papageorghiou AT, Parra M, Palma Dias R, Nicolaides KH; Fetal Medicine Foundation Second Trimester Screening Group. Randomized controlled trial using low-dose aspirin in the prevention of pre-eclampsia in women with abnormal uterine artery Doppler at 23 weeks’ gestation. Ultrasound Obstet Gynecol. 2003 Sep;22(3):233–9. doi: 10.1002/uog.218. PMID: 12942493.

